# Treatment seeking behaviours, antibiotic use and relationships to multi-drug resistance: A study of urinary tract infection patients in Kenya, Tanzania and Uganda

**DOI:** 10.1101/2023.03.04.23286801

**Authors:** Keina Sado, Katherine Keenan, Areti Manataki, Mike Kesby, Martha F Mushi, Stephen E Mshana, Joseph Mwanga, Stella Neema, Benon Asiimwe, Joel Bazira, John Kiiru, Dominique L Green, Xuejia Ke, Antonio Maldonado-Barragán, Mary Abed Al Ahad, Kathryn Fredricks, Stephen H Gillespie, Wilber Sabiiti, Blandina T Mmbaga, Gibson Kibiki, David Aanensen, V Anne Smith, Alison Sandeman, Derek J Sloan, Matthew TG Holden, HATUA Consortium

**Affiliations:** University of St Andrews, St Andrews, UK; Catholic University Of Health And Allied Sciences, Mwanza, Tanzania; Makerere University, Kampala, Uganda; Mbarara University of Science and Technology, Mbarara, Uganda; Kenya Medical Research Institute, Nairobi, Kenya; Kilimanjaro Clinical Research Institute, Kilimanjaro Christian Medical Centre, Moshi, Tanzania; Kilimanjaro Christian Medical University College, Moshi Tanzania; Africa Excellence Research Fund, London, UK; Oxford Big Data Institute, Oxford, UK; London School of Hygiene and Tropical Medicine, London, UK

**Keywords:** healthcare use, antibiotic use, antibacterial resistance, Sub-Saharan Africa, Urinary tract infection

## Abstract

Antibacterial resistance (ABR) is a major public health threat. An important accelerating factor is treatment-seeking behaviours, including inappropriate antibiotic (AB) use. In many low- and middle-income countries (LMICs) this includes taking ABs with and without prescription sourced from various providers, including health facilities and community drug sellers. However, investigations of complex treatment-seeking, AB use and drug resistance in LMICs are scarce.

The Holistic Approach to Unravel Antibacterial Resistance in East Africa (HATUA) Consortium collected questionnaire and microbiological data from 6,827 adult outpatients with urinary tract infection (UTI)-like symptoms presenting at healthcare facilities in Kenya, Tanzania and Uganda. Among 6,388 patients we analysed patterns of self-reported treatment seeking behaviours (‘patient pathways’) using process mining and single-channel sequence analysis. Of those with microbiologically confirmed UTI (n=1,946), we used logistic regression to assessed the relationship between treatment seeking behaviour, AB use, and likelihood of having a multi-drug resistant (MDR) UTI.

The most common treatment pathways for UTI-like symptoms included attending health facilities, rather than other providers (e.g. drug sellers). Patients from the sites sampled in Tanzania and Uganda, where prevalence of MDR UTI was over 50%, were more likely to report treatment failures, and have repeated visits to clinics/other providers, than those from Kenyan sites, where MDR UTI rates were lower (33%). There was no strong or consistent relationship between individual AB use and risk of MDR UTI, after accounting for country context.

The results highlight challenges East African patients face in accessing effective UTI treatment. These challenges increase where rates of MDR UTI are higher, suggesting a reinforcing circle of failed treatment attempts and sustained selection for drug resistance. Whilst individual behaviours may contribute to the risk of MDR UTI, our data show that factors related to context are stronger drivers of ABR.

## Introduction

Antibacterial resistance (ABR) is a major global health challenge which compromises our ability to treat infections with antibiotics (ABs)(1). ABR was associated with an estimated 5 million deaths globally in 2019, and the annual mortality rate from ABR is expected to increase further by 2050(2,3). The burden is greatest in low- and middle-income countries (LMICs)(2), which experience a higher prevalence of infectious diseases, and have fewer technical and financial resources to deal with multi-drug resistant (MDR) pathogens. Whilst the genetic mutations which make microbes resistant to ABs occur spontaneously(4), overuse and misuse of ABs to treat human illness is a major driver of ABR in pathogens that cause disease(5). Many studies have identified increased AB use, and prior AB use, as risk factors for ABR infections, driving resistance through exerting bacterial selection pressures within individuals and communities(6–9). Deeper understanding of the dynamics of treatment-seeking, AB use and ABR in LMIC settings is therefore crucial to tackle the problem. Untangling cause and effect between high AB use and ABR is challenging, especially where ABR infections are more common, and repeat AB use may be a reaction to treatment failure.

Healthcare and treatment landscapes in LMICs are sometimes described as ‘pluralistic’(10), which typically means there are multiple sources of public and private clinics, alongside pharmacists, drug sellers, and complementary and traditional/herbal medicine providers. This multi-layered context of care provision overlaps with models of medical syncretism, which describe how patients make treatment decisions drawing on hybrid logics(11), composed of both biomedical understandings and local knowledge/beliefs present in different contexts(11,12). Decision-making in LMICs is further influenced by a number of individual and structural factors, including under-resourced public health provision, stock- outs of medicines and multi-layered systems of healthcare insurance(13–15). This potentially leads to great diversity in pathways to AB use among patients, something which has been recently highlighted as a research gap(10).

Existing research suggests that AB self-medication from drug shops is very common in parts of Asia and Sub-Saharan Africa(14–18). This is facilitated, in some areas, by the ease of purchasing ABs without prescription at community pharmacies and drug shops(19), and the possibility of buying partial courses which can result in sub-optimal treatment dosing.

Studies also suggest that consumption of leftover ABs (i.e. previously obtained but unconsumed) is common(17). Thus far, there have been no studies investigating the interrelationship between patient AB use and ABR in East Africa, a region with high levels of ABR(2,20), high prevalence of non-prescription drug sales(21) and complex, hybrid treatment landscapes.

In previous studies, AB use or ‘misuse’ has often been measured simply as, for example, a binary indicator of any type of self-medication(16). However, situating treatment-seeking behaviours in their appropriate social and temporal context is vital. AB self-medication may occur as part of a longer sequence of treatment attempts(22), or in parallel to other attempts, emphasising the importance of a longitudinal, pathway based approach(23).

Pathway analysis has been used to investigate care for tuberculosis,(24) abortion,(25) and cancer(26) but has rarely been used to understand AB use (15). Appropriate data and methods are needed to capture the behavioural nuance inherent in such patient pathways. This study makes use of exceptionally detailed linked quantitative social and microbiological data collected as part of the part of a multi-country interdisciplinary consortium “Holistic Approach to Unravel Antibacterial Resistance in East Africa (HATUA)” which aimed to delineate the drivers of antibiotic resistance in this community.(27). The study focused on urinary tract infections (UTIs) as a lens for understanding treatment-seeking and ABR more generally. UTIs are globally common bacterial illnesses which are usually treated with ABs, but can cause threatening complications if ineffectively treated in vulnerable patients(28,29).

In this study, we address two research questions: 1) What are the main characteristics of treatment-seeking pathways among patients with UTI-like symptoms in Kenya, Tanzania, and Uganda? and 2) How do treatment-seeking behaviours and AB use relate to the likelihood of developing an UTI caused by a multi-drug resistant (MDR) pathogen (i.e. resistant to at least 3 classes of antibiotic)?

## Materials and Methods

### Data and sample

The original sample consisted of 6,827 adult outpatients with UTI-like symptoms (e.g. burning when urinating, frequent urination, blood or pus in urine), aged 18 years and over , and pregnant adolescents aged 14-17 years, who were recruited from three sites in Kenya (Makueni, Nairobi, and Nanyuki), Tanzania (Kilimanjaro, Mbeya, and Mwanza), and Uganda (Mbarara, Nakapiripirit, and Nakasongola), between February 2019 and September 2020 (see Figure 1). We recruited patients at medical facilities that were predominantly government-funded (see Appendix Table S1 for a full explanation). In all sites, less than 1% of those approached declined to participate. Ethical approval was obtained from National and Institutional Research Ethics Committees (see protocol(27)).

**Figure 1.**
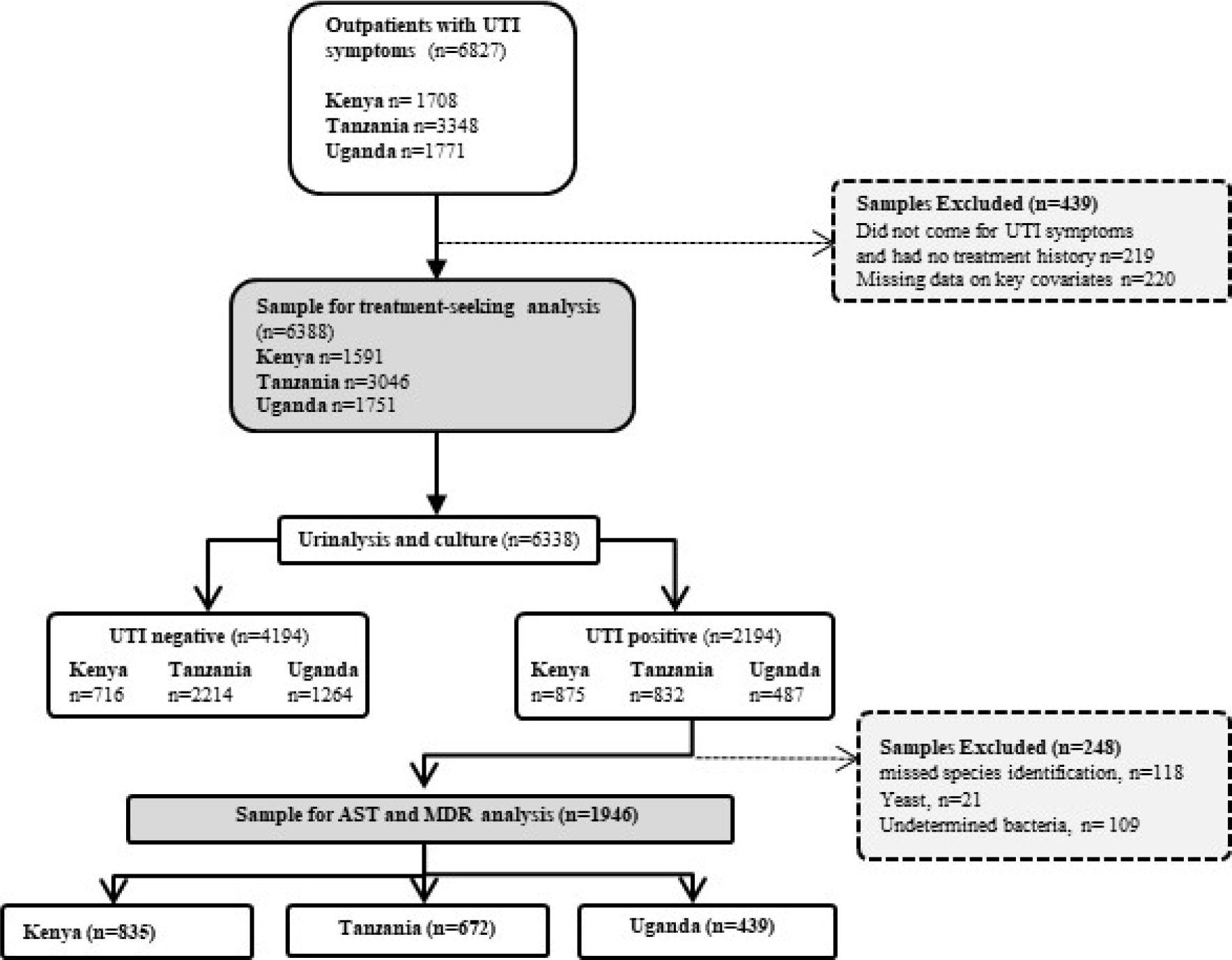
Selection of the analysis samples

**Table 1:**
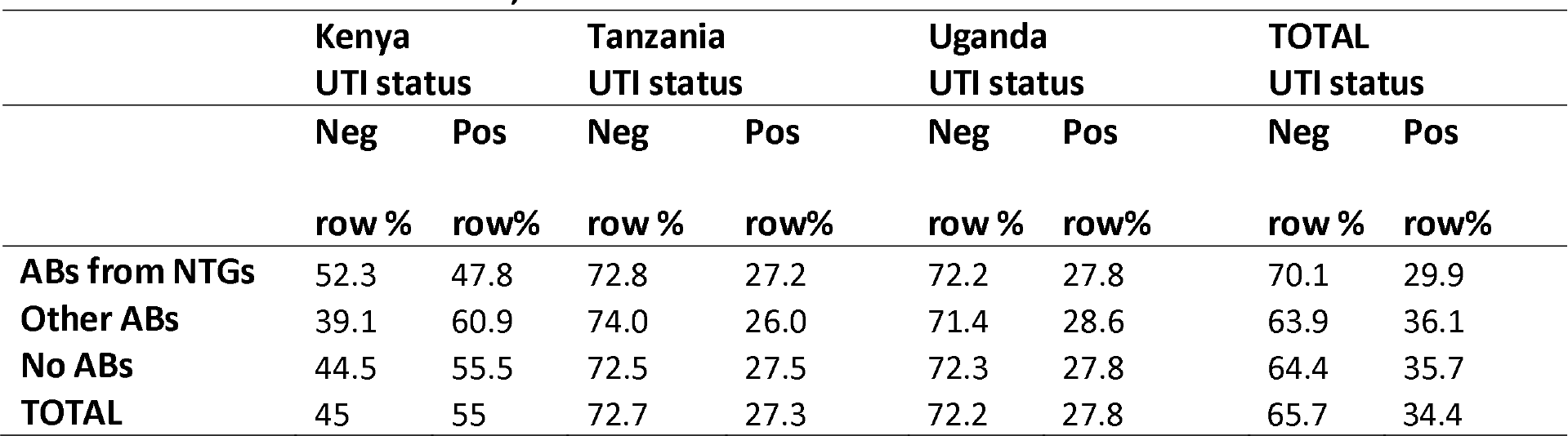
AB use and UTI status, across 3 countries

### Microbiological identification of UTI and antibiotic sensitivity testing (AST)

All patients provided a clean catch mid-stream urine sample, which underwent microbiological culture, pathogen identification and AST. Urinary tract infection positive samples (UTI+) were defined by the presence of >10^4^ colony-forming units per millilitre (CFU/mL) of one or two uropathogens. For UTI positive patients, susceptibility to the tested antibiotics (see appendix Table S2 for a full list) was done by the disk diffusion method, and determined using breakpoints (zone diameter interpretive criteria) indicated in the 2021 CLSI guidelines (M02 document) (30) Multidrug-resistant (MDR) bacteria were defined as urinary isolates resistant to at least one agent in three or more categories of antimicrobial agents, following the European Centre for Disease Prevention and Control (ECDC) guidelines(31) with some modifications. Specifically, nitrofurantoin and trimethoprim, two antibiotics routinely used for treating UTIs that are not included in the ECDC guidelines, were also considered for estimating MDR. In addition, for those species/genera not incorporated in the ECDC guidelines, i.e. *Salmonella, Shigella* and *Streptococcus,* the MDR rates were calculated as above, but considering the resistance to a selected pool of tested antibiotics (Table S2). Isolates that showed intermediate resistance to a given antibiotic were considered resistant. .

**Table 2:** Sociodemographic, contextual and behavioural factors and their bivariate association with MDR

| | | MDR-<br>n(%) | MDR+<br>n(%) | $\chi^2$, p value |
| --- | --- | --- | --- | --- |
| **Country** | Kenya | 553 (66.2) | 282 (33.8) | 117.6 (p<0.001) |
|  | Tanzania | 272 (40.5) | 400 (59.5) |  |
|  | Uganda | 189 (43.1) | 250 (56.9) |  |
| **Gender** | Male | 115 (41.5) | 162 (58.5) | 14.0 (p<0.001) |
|  | Female | 899 (53.9) | 770 (46.1) |  |
| **Age** | <25 | 298 (56.7) | 228 (43.3) | 27.3 (p<0.001) |
|  | 25-34 | 372 (55.3) | 301 (44.7) |  |
|  | 35-44 | 152 (52.6) | 137 (47.4) |  |
|  | 45-54 | 77 (44.0) | 98 (56.0) |  |
|  | 55-64 | 33 (37.1) | 56 (62.9) |  |
|  | 65+ | 82 (42.3) | 112 (57.7) |  |
| **Education** | None | 131 (46.5) | 151 (53.5) | 56.4 (p<0.001) |
|  | Primary | 280 (42.6) | 378 (57.4) |  |
|  | Secondary | 383 (57.7) | 281 (42.3) |  |
|  | Higher | 220 (64.3) | 122 (35.7) |  |
| **Delay** | Less than 2 weeks | 817 (55.1) | 666 (44.9) | 21.7 (p<0.001) |
|  | More than 2 weeks | 197 (42.5) | 266 (57.5) |  |
| **Treatment steps** | 1(straight to clinic) | 557 (56.5) | 428 (43.5) | 27.9 (p<0.001) |
|  | 2 | 292 (52.3) | 266 (47.7) |  |
|  | 3+ | 165 (40.9) | 238 (59.1) |  |
| **First care accessed** | Recruitment clinic | 557 (56.5) | 428 (43.5) | 20.1 (p<0.001) |
|  | Clinic | 309 (45.5) | 370 (54.5) |  |
|  | Pharmacy/drug shop | 79 (54.5) | 66 (45.5) |  |
|  | Self-treatment | 69 (50.4) | 68 (49.6) |  |
| **Any AB use in pathway** | No | 781 (54.1) | 663 (45.9) | 8.5 (p=0.004) |
|  | Yes | 233 (46.4) | 269 (53.6) |  |
| **Any AB use in past 6 months** | No | 425 (52.9) | 379 (47.1) | 0.3 (p=0.608) |
|  | Yes | 589 (51.6) | 553 (48.4) |  |

### Variables: treatment-seeking behaviours

Patients answered a questionnaire on treatment-seeking for UTI symptoms, previous AB use practices and attitudes, and sociodemographic characteristics. Figure 2 illustrates the structured questionnaire used to collect treatment-seeking data, which identified the types of providers consulted, treatments/ABs taken, and reasons for these choices prior to attending the recruitment clinic. We derived a variable for the number of steps in the pathway (1 step, i.e. came straight to recruitment clinic, 2 steps, i.e. took one additional step before coming to recruitment clinic, and 3 or more steps i.e. took 2 or more steps before being recruited). During the interview, patients self-reported the names of medicines they had taken to treat their UTI-like symptoms, and were also prompted using a drug bag or drug card developed specifically for each site(32). Subsequently, we assessed whether the AB they reported was recommended for treating UTI in adult outpatients according to National Treatment Guidelines (NTGs) for their country (see appendix Table S3).

**Figure 2:**
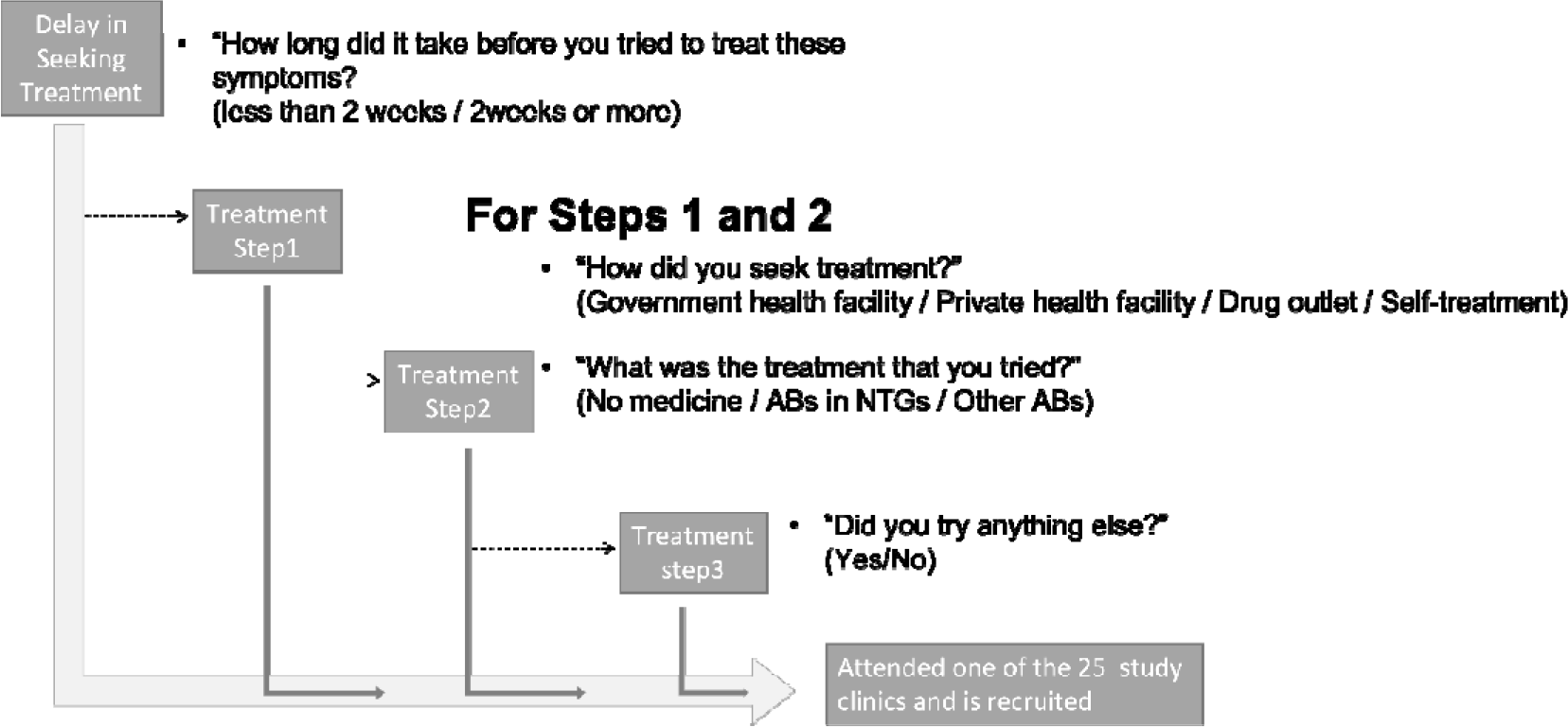
Treatment-seeking questions from the HATUA questionnaire and derived outcomes in this study

**Table 3:** Odds ratios for multi-drug resistant UTI pathogens according to treatment seeking, sociodemographic factors and country

|  | Kenya (n=835) |  | Tanzania (n=672) |  | Uganda (n=439) |  |
| --- | --- | --- | --- | --- | --- | --- |
|  | Model 1 | Model 2 | Model 1 | Model 2 | Model 1 | Model 2 |
|  | OR (95% CI) | OR (95% CI) | OR (95% CI) | OR (95% CI) | OR (95% CI) | OR (95% CI) |
| **Treatment** |  |  |  |  |  |  |
| sequence (ref:A) |  |  |  |  |  |  |
| B | 1.17(0.69-2.00) | 1.09(0.63-1.87) | 0.94(0.54-1.66) | 0.87(0.49-1.56) | **2.06(1.01-4.19)** | 1.66(0.79-3.51) |
| C | 0.72(0.25-2.06) | 0.73(0.25-2.09) | 1.11(0.56-2.17) | 1.07(0.54-2.14) | **2.29(1.09-4.8)** | 1.72(0.78-3.8) |
| D | 0.60(0.27-1.36) | 0.5(0.21-1.16) | 0.85(0.41-1.74) | 0.8(0.38-1.7) | 1.45(0.73-2.86) | 1.27(0.63-2.57) |
| E | 0.7(0.40-1.25) | 0.64(0.36-1.15) | 0.91(0.5-1.67) | 0.86(0.46-1.59) | 1.55(0.77-3.14) | 1.31(0.62-2.76) |
| F | 1.12(0.73-1.72) | 1.12(0.72-1.74) | 0.65(0.31-1.36) | 0.59(0.28-1.25) | 2.18(0.95-4.99) | 2.06(0.88-4.79) |
| G | 1.34(0.42-4.28) | 1.23(0.37-4.04) | 0.89(0.53-1.5) | 0.83(0.48-1.41) | 0.9(0.38-2.11) | 0.64(0.25-1.6) |
| H | 0.47(0.13-1.68) | 0.45(0.12-1.66) | 1.48(0.83-2.64) | 1.45(0.8-2.65) | 1.76(0.82-3.8) | 1.3(0.56-2.99) |
| I + J combined | 0.21(0.03-1.66) | 0.16(0.02-1.32) | 1.18(0.69-2.04) | 1.05(0.59-1.88) | **3.94(1.76-8.86)** | **2.86(1.2-6.83)** |
| **Gender** |  |  |  |  |  |  |
| (ref:Female) |  |  |  |  |  |  |
| Male |  | 0.87(0.5-1.53) |  | **0.53(0.33-0.85)** |  | 1.44(0.77-2.67) |
| **Age (ref:<25)** |  |  |  |  |  |  |
| 25-34 |  | 1.29(0.91-1.83) |  | 1.23(0.75-2) |  | 1.14(0.7-1.85) |
| 35-44 |  | 1.31(0.81-2.13) |  | 0.73(0.43-1.26) |  | 1.42(0.74-2.71) |
| 45-54 |  | **2.24(1.09-4.6)** |  | 0.78(0.43-1.43) |  | 1.36(0.7-2.66) |
| 55-64 |  | 1.83(0.52-6.5) |  | 1.2(0.62-2.29) |  | 1.66(0.49-5.62) |
| 65+ |  | 1.34(0.53-3.37) |  | 0.83(0.46-1.49) |  | 0.89(0.29-2.71) |
| **Education** |  |  |  |  |  |  |
| (ref:Higher) |  |  |  |  |  |  |
| None |  | 1.09(0.28-4.23) |  | 0.83(0.53-1.32) |  | **1.75(1.03-2.99)** |
| Primary |  | 0.95(0.25-3.59) |  | 0.84(0.48-1.5) |  | 1.15(0.62-2.11) |
| Secondary+ |  | 0.76(0.2-2.91) |  | 0.5(0.22-1.11) |  | 1.63(0.75-3.56) |

NB. Bold indicates estimates where 95% CIs do not cross 1.

### Other variables

We included patients’ self-reported sex (male/female), age (categorised into <25; 25-34; 35- 44; 45-54; 55-64; 65+ years), and education level (none, primary, secondary, and tertiary).

We also included a binary variable measuring use of any other ABs (apart from those reported in the pathway) in the previous 6 months.

### Analytical Approach

#### Characterising treatment seeking behaviours

We employed computational methods of process mining and sequence analysis that are designed to detect patterns in process /flow data. The application of these methods to clinical healthcare data in LMICs is novel. Process mining is typically used to analyse event sequences automatically recorded in high-income settings, such as electronic inpatient records(30). Sequence analysis is more commonly applied in biological genetics and sociology(33), but use to analyse healthcare processes is being developed(34).

#### Process mining

Self-reported responses shown in Figure 2 were organised in a temporal sequence for each patient, capturing delay (or not)-> step 1->treatment at step 1-> step 2->treatment at step 2->step 3. Patients who arrived at clinic at an earlier stage had shorter sequences. We assigned timestamps that reflected temporal sequences of individual patients. We also merged some categories of variables due to low frequency. For example, for the question “How did you seek treatment?”, we merged responses “Visiting government health facility” and “Visiting private health facility” into “Visiting health facility”. We employed the Heuristic Miner using the PM4Py package in Python (35). The process is visually described in Figure 3. The output of the heuristic miner is a process model that captures the control-flow relations between tasks that are observed in the data, including the number of patients that moved from one step to another. In our study, the process model provides a general overview of all treatment seeking pathways. Subsequently, we visualised the most common pathways by analysing traces (i.e. sequences of steps) with the use of ProM(36) software.. We compared the numbers/proportions of patients taking common trajectories and qualitatively compared the traces across countries.

**Figure 3.**
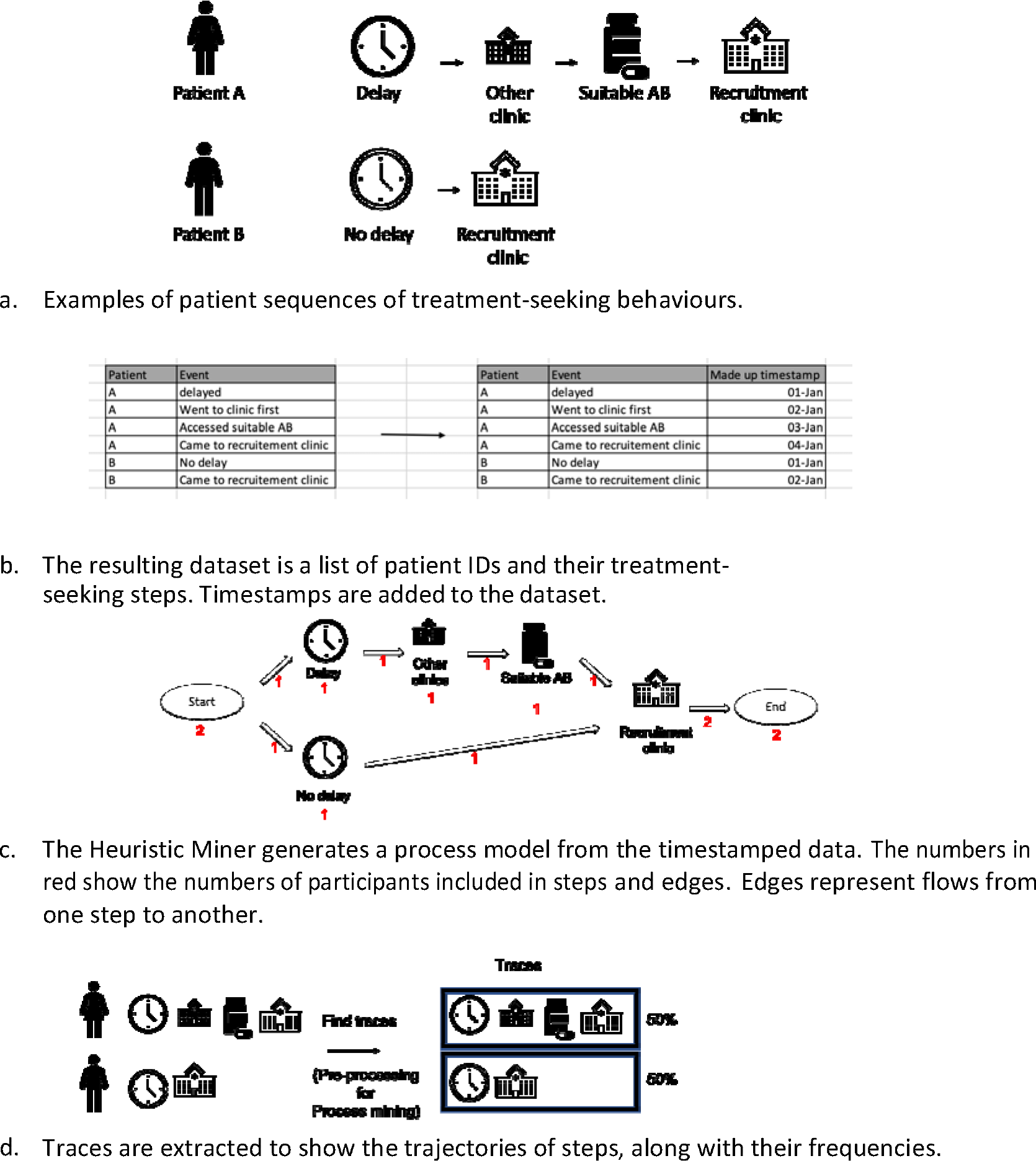
**A-D**S:tages in data management, analysis and visualizationfor process mining

Next, we used **single-channel sequence analysis** (WeightedCluster library in R(37)) to cluster patients with similar treatment-seeking pathways. To derive the clusters, we used the variables shown in Figure 2, subsequently recoded, as follows: (delay (yes/no), types of provider at step 1 and 2 (health facility/drug outlet/self-treatment), treatment at step 1 and 2 (ABs from NTGs/ other ABs/ no ABs), and number of steps (1,2,3+). In the preliminary stages of analysis, we noticed that the clustering was strongly driven by the number of steps. Therefore, to allow us to investigate other aspects of treatment seeking, we repeated the clustering exercise separately among those who took 1 step, 2 steps, and 3 or more steps. Following others(38), we used the optimal matching algorithm to calculate dissimilarity measures. Because all sequences in each subgroup have the same number of steps, costs for insertion/deletion were not required. Substitution costs were set to a constant value (the default). The optimal number of clusters were evaluated using two cluster quality measures: Point Biserial Correlation (PBC), and Weighted Average Silhouette width (ASWw)(37). We assessed the characteristics of cluster membership by conducting a bivariate analysis assessing the associations between age, gender, education, country and cluster membership, using chi square tests for difference.

#### Associations between social factors, behaviours and MDR status

To investigate associations between treatment seeking and MDR, we used a subset of UTI positive patients with valid, linked data on MDR, treatment-seeking and socio-demographic characteristics (n=1,946). We first cross-tabulated separate facets of treatment-seeking characteristics (delay, types of provider at every step, treatments at every step, number of steps) and socio-demographics (age, gender, education, country) with MDR status, and conducted chi-square tests for difference. Then, we assessed the association between cluster membership (derived from the sequence analysis described above) and MDR status using multivariable logistic regression, stratified by country.

## Results

### Sample description

To analyse patterns in UTI-treatment-seeking pathways, we excluded 219 patients who came to the recruitment clinic for non-UTI symptoms and had not attempted to treat their symptoms, leaving n= 6,608 patients. After further exclusions for missing covariate data (n=220), the remaining 6,388 patients were composed of 1,591 (24.9%), 3,046 (47.7%), 1,751 (27.4%) from Kenya, Tanzania and Uganda respectively (see Figure 1). Table S4 shows the characteristics of patients included in the two different analysis samples (stratified by country in Tables S5-7). Considering the sample used for pathway analysis (n=6,338), nearly half of the patients were recruited in Tanzania (47.7%), most were female, and of reproductive age (<44 years). Overall, most patients had either primary or secondary level education, with much smaller proportions with no or higher education. In terms of treatment seeking pathways, while many patients (45.1%) had come straight to the recruitment clinic, the majority (54.9%) had taken at least one additional treatment step prior to recruitment into the HATUA study. Approximately 30% of patients had already taken ABs to treat their UTI-like symptoms, and over 60% reported having taken ABs in the previous 6 months. Of the total patients, 34.3% of participants had a UTI pathogen microbiologically confirmed, and of these, nearly half (47.9%) had an MDR pathogen responsible for their infection.

To analyse associations with MDR (shaded green box in Figure 1), we further restricted the sample to microbiologically confirmed UTI patients with AST data for the pathogen identified. We excluded patients if the pathogen or causative bacteria was unknown, or where a breakpoint for disk diffusion test was unavailable. This resulted in a sample of n=1946 which had slightly different distributions to the sample of all patients (Table S4-S7). For example, a higher proportion came from Kenya and fewer from Tanzania, and women and older people made up a larger fraction of the sample. This is related to the higher rates of confirmed UTI in those different subgroups influencing their eventual inclusion into the sample with MDR data.

### Characteristics of treatment-seeking pathways

Figure 4 visualizes the treatment-seeking pathways derived from process mining for all countries combined. The majority (72%) of our patients accessed care within 2 weeks of recognizing their symptoms, while 28% waited longer than 2 weeks. The most common first step was visiting one of the study recruitment clinics (46% of patients), and the second most common was visiting other health facilities (41%), followed by visiting drug outlets (7%) and self-treatment (7%). At every step, attending health facilities, either the recruitment clinic or another government-funded facility, were the predominant steps (it is important to emphasize that our clinic-based recruitment strategy has likely led to bias in this direction). Finally, 644 (10% of all participants) tried three or more things before visiting the recruitment clinic. Among patients who did something before visiting the recruitment clinic (3,509 patients), at the first step, 39% took ABs recommended in the NTGs for UTI, 9% took other ABs, and 52% took other medications or nothing at all. At the second step (n=1,674), there were similar proportions (33% took ABs in NTGs, 8% took other ABs).

**Figure 4:**
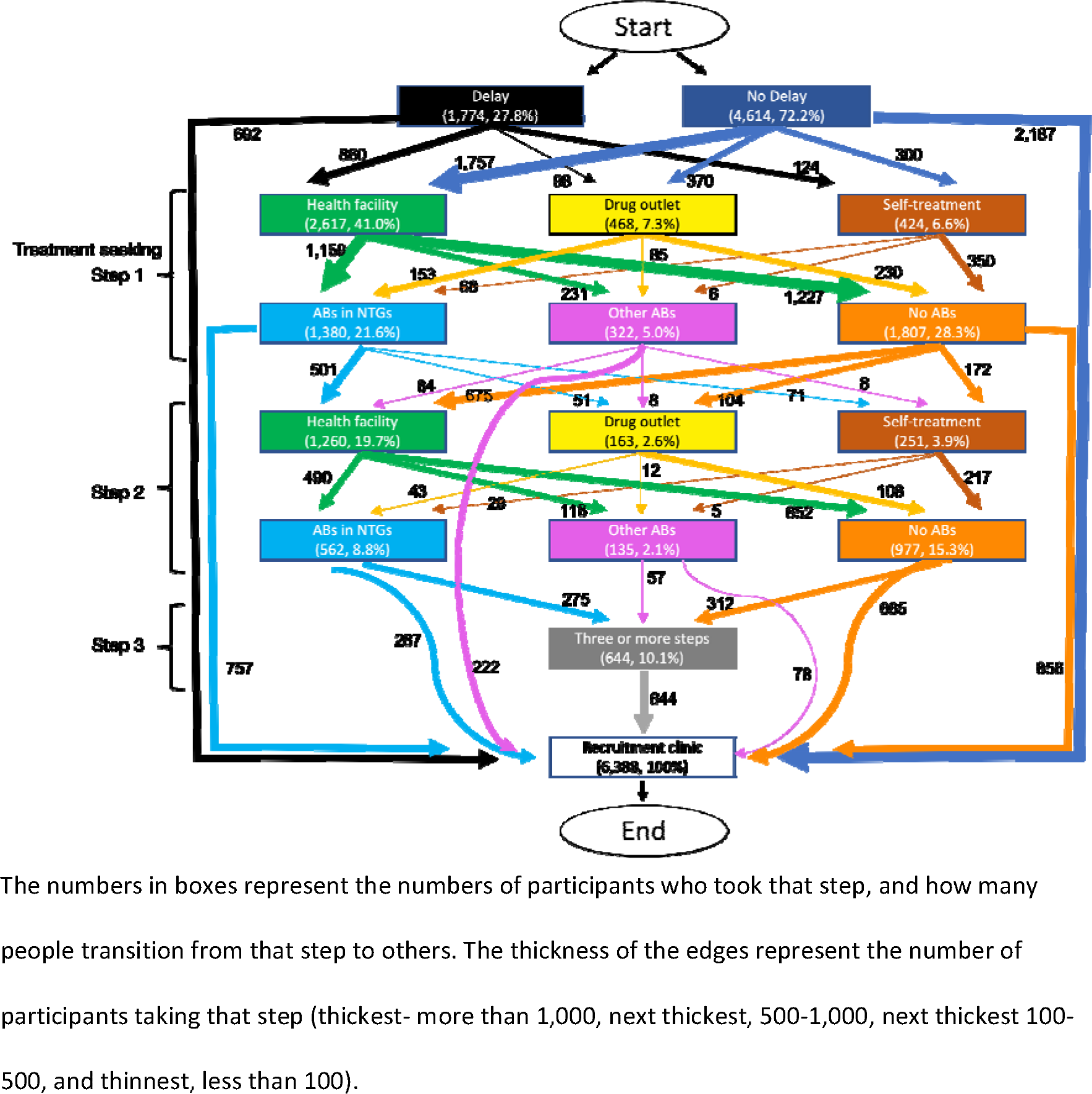
Treatment seeking pathways visualized with heuristic miner among patients in Kenya, Tanzania and Uganda. The numbers in boxes represent the numbers of participants who took that step, and how many people transition from that step to others. The thickness of the edges represent the number of participants taking that step (thickest- more than 1,000, next thickest, 500-1,000, next thickest 100- 500, and thinnest, less than 100).

Using the Trace Explorer function, we detected 183 unique treatment seeking patterns. Logically, shorter pathways were the most common because there was less opportunity for variation. The most frequently taken pathways were visiting the recruitment clinic either within 2 weeks of noticing symptoms (34% of patients), or after 2 weeks (11%). Visiting other types of clinics (government or private) and taking no medication or ABs in NTGs represented the third, fourth and fifth most-common pathways. It was only at the sixth most common when non-healthcare facility based treatments emerged. The majority of treatments taken were either ABs in NTGs, or other medicines (taking other ABs was the least common type of treatment).

We considered country-level differences in the 10 most common pathways (see Figure 5) and this showed that Kenyan patients were more likely to access care without delay and directly at the recruitment clinic than those in Tanzania or Uganda (55% of Kenyan patients compared with 24% in Tanzania and 33% in Uganda). On the other hand, Kenyan patients who did not go straight to recruitment clinic were the most likely to try self-treatment (5%) or visit a drug outlet (4%) first. Among Tanzanian patients, the most common trajectories all featured healthcare facilities visits, but patients were more likely to have 2 and 3 step pathways than other countries (>10% of all sequences). Ugandan patients had simpler and more healthcare-dependent pathways than in Tanzania.

**Figure 5:**
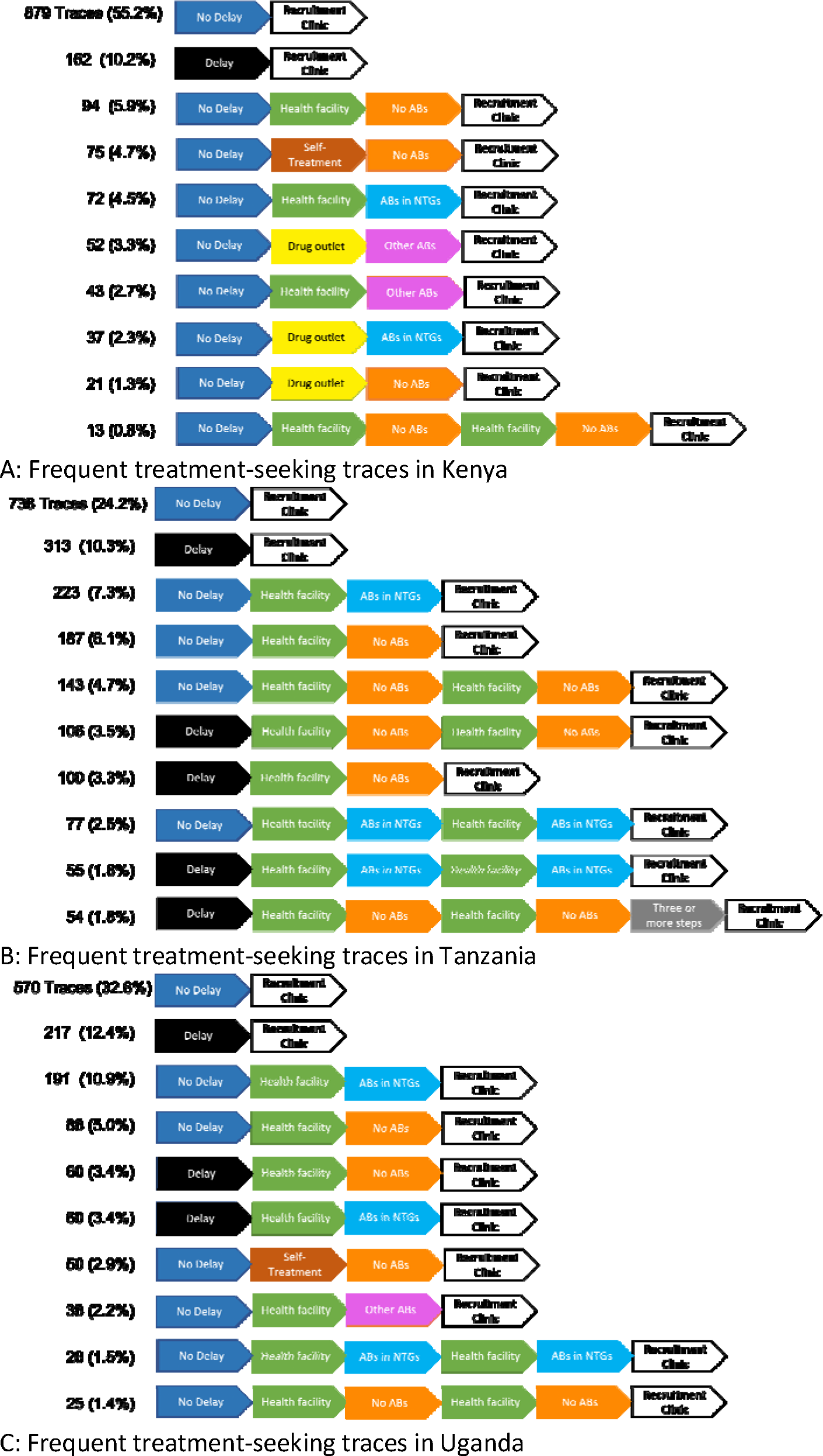
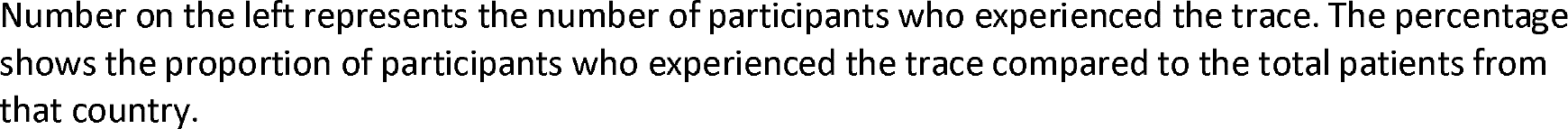
Ten most common treatment-seeking patterns visualized in Kenya (panel A), Tanzania (panel B) and Uganda (panel C). Number on the left represents the number of participants who experienced the trace. The percentage shows the proportion of participants who experienced the trace compared to the total patients from that country.

Figure 6 shows the total distribution of patients taking any AB courses during the pathway, whether they were in the NTGs for UTI or not, and whether they were taken after a visit to healthcare facilities, drug outlets, or other. Overall, taking any type of ABs was less likely to occur in Kenya than in Tanzania or Uganda ( a finding driven by the generally shorter pathways Kenyan patients reported). However, Kenyan patients had the highest rate of taking ABs not in the NTGs, and were more likely to access these from drug outlets, than in the other countries. In all countries, taking ABs was most likely to happen after a visit to a healthcare facility. Kenya patients were proportionally more likely to visit pharmacies/drug shops prior to taking ABs, than in Tanzania or Uganda. Patients with no microbiologically confirmed UTI were just as likely to have taken ABs to treat their UTI-like symptoms as those with a UTI confirmed (see Table 1). Of all the patients who took ABs, two thirds (66%) were patients with no confirmed UTI.

**Figure 6:**
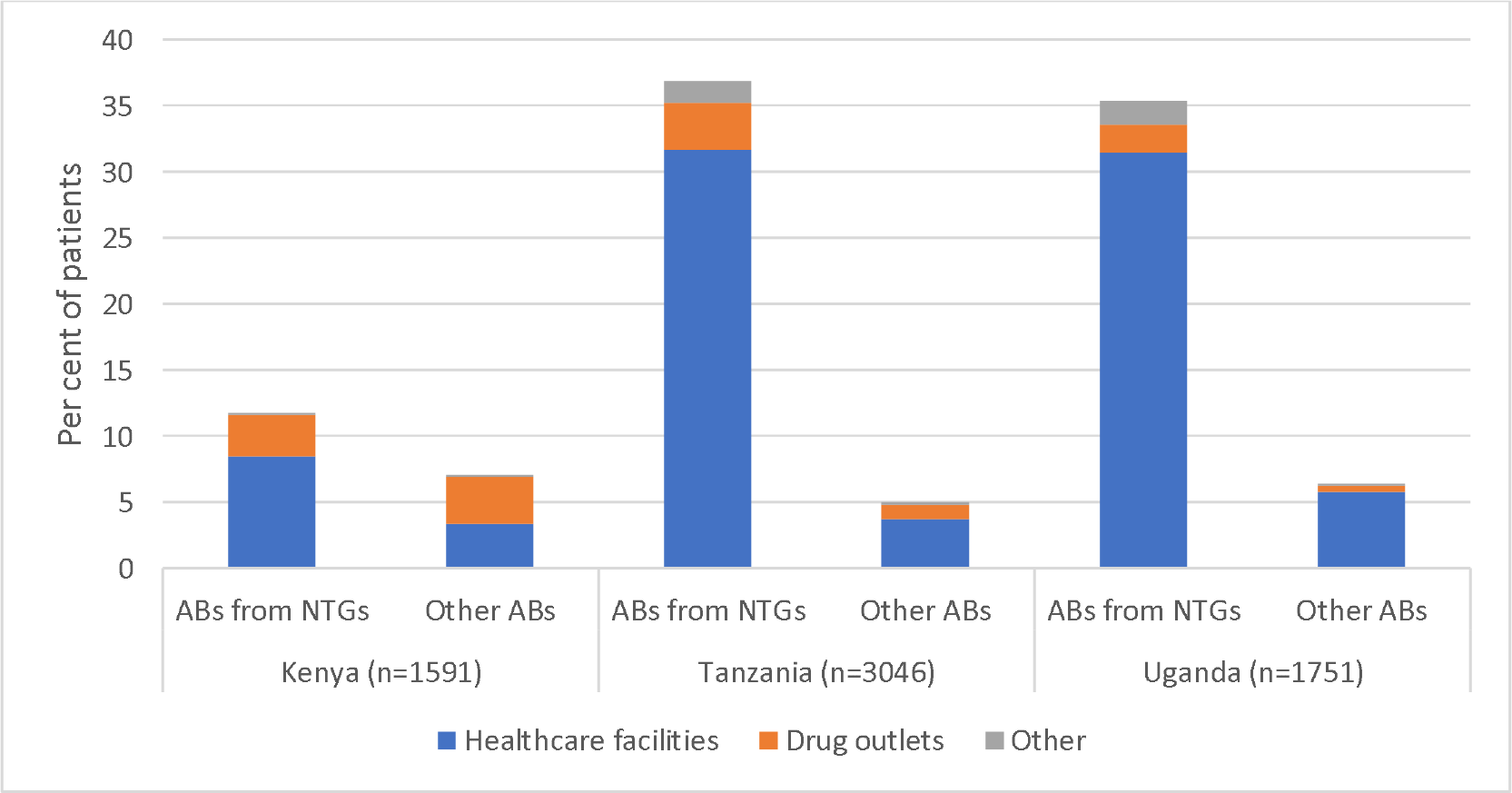
Percentage of patients taking ABs for their UTI-like symptoms at any point prior to being recruited, by country and type of provider.

### Clustering of treatment-seeking behaviours

After dividing the data into three subgroups: 1-step, 2-step, and 3-step or longer (3+ steps) sequences, we conducted sequence analysis separately on each subgroup. The cluster quality measures are shown in Figures S1 and S2. Since no strong peaks were observed in these measures, the final number of clusters was chosen as a trade-off between having a reasonably low number to maximize sample size for the subsequent analysis, with reasonably high scores on quality measures. Using this approach 10 clusters were detected: 2 clusters from the 1-step sequence subgroup, and 4 clusters each from the 2-steps and 3+ steps subgroups. Figure 4 summarizes the distribution of behavioural characteristics of these clusters by country, gender, education, and age. The characteristics of the clusters are further visualized in Supplementary Material Figure S3.

As might be expected from the process mining analysis, Kenyan patients were less likely to belong to more complex pathways than those from Tanzania or Uganda. They were also proportionally more likely to belong to cluster F (associated with visiting drug outlets, and taking ABs not in NTGs). Tanzanian patients were the most likely to take 3 or 3+ step pathways. Higher proportions of males belonged to the clusters G-J, indicating they were more likely to have complex pathways. The pathways showing the highest mean age were also the longest (I & J). There did not seem to be a clear relationship between education and cluster membership.

### Associations between treatment-seeking, AB use and MDR

The next stage of the analysis uses a smaller subset of patients with complete data on MDR (n=1,946). Patients excluded due at this stage were more likely to be from Tanzania or Uganda, to be older, male, and have primary education (Table S8). Table 2 shows bivariate associations between patient characteristics and MDR status. Patients with higher levels of MDR were more likely to be from Uganda and Tanzania than Kenya; to be male, older, and have no studies or primary school education only. Patients with MDR infections are also more likely to have reported delay in accessing care; 3+ treatment-seeking steps; accessed a healthcare facility as their first step; and using ABs in their pathway. Notably, AB use in the previous 6 months was not associated with higher MDR rates. We repeated the bivariate analysis with MDR stratifying on country (see Table S9). Some relationships held across settings, but others did not. For example, in Kenya and Tanzania, males have higher levels of MDR, but the gender relationship was reversed in Uganda. Middle-aged individuals (either 45-54 or 55-64 years) had the highest levels of MDR across all 3 countries. There was no consistent educational gradient. The association between delay in seeking treatment, higher treatment steps, more AB use, and MDR was more apparent in Tanzania and Uganda than Kenya.

Due to there being strong country differences in the distribution of independent and dependent variables, we investigated the relationship between pathway clusters and MDR using multivariate logistic regression in country-specific models (Table 3). In Kenya and Tanzania there is no association between pathway clusters and risk of MDR. In Uganda, clusters B, C, and I-J have significantly higher odds of MDR than those in cluster A. After controlling for age, gender and education (model 2), those in clusters I-J (3-step pathways) still had significantly higher odds of MDR. In models combining all three countries, there is no association between treatment pathways, individual factors and MDR once country is adjusted for.

## Discussion

This study used process mining and sequence analysis to characterise patterns of treatment-seeking behaviour among adult UTI outpatients in Kenya, Tanzania, and Uganda, and to investigate correlations between those behavioural patterns and MDR UTIs. Consistent with previous work (13), in all three countries behaviour was dominated by visits to healthcare facilities. However, over half of patients had apparent treatment failures, leading to repeat clinic visits within the same UTI episode. Repeat clinic visits were more common in Tanzania and Uganda and were associated with higher rates of AB use (regardless of their suitability to treat UTI). While Kenyan patients typically had simpler pathways, they were also more likely to consult with pharmacists and drug sellers, and to take ABs not recommended for UTI in the Kenyan National Treatment Guidelines. The relationship between health behaviours and MDR rates depended on context. In Uganda, where over half of UTI patients had MDR pathogens, those with longer sequences of treatment steps had higher rates of MDR UTI, but this was not the case in Kenya (where MDR prevalence was lower, at 33%).

Thus, the main message is that individuals with complex treatment seeking pathways, and repeat clinic visits, are also more likely to have had higher AB use. Past treatment behaviours, however, are not consistently correlated with higher risk of MDR UTI at the individual level, suggesting that other contextual factors contribute to MDR risk.

The previous (predominantly qualitative) literature on the African healthcare landscape and patient pathways has emphasised the plurality of treatment seeking behaviours, high prevalence of self-treatment, diversity of treatment providers and parallel systems of allopathic and traditional/herbal medicine(11,39–41). Our study presents a different picture, where the majority behaviour in all three countries was to seek help with formal medical facilities. This is likely due to the sampling bias inherent in our study design. By recruiting clinic attendees, we select those who are most likely to visit those facilities, and miss those who seek self-treatment. Nevertheless, despite this acknowledged bias, we argue that the results highlight an often overlooked truth about the drivers of ABR in LMICs. That is, that one of the most important complexities experienced by patients in this healthcare landscape arises from difficulties in getting effective care for infections within the formal healthcare system. Thus, common infections are not necessarily simple to treat or easy to resolve. This basic challenge may promote medical pluralism out of necessity, and results in more costs for patients and the healthcare system, more medicines taken, and more suffering. This challenge is undoubtedly exacerbated further still by the developing AMR pandemic, which make those apparently ‘simple’ infections difficult to treat.

A further striking finding is the high proportion of patients who ostensibly take ABs to treat UTI-like symptoms,but turn out to not have a UTI infection. Patients without a UTI pathogen were equally likely to take ABs as those who had one, and given the UTI posivity rate in the study, this means that two thirds of our recruited patients take ABs for a UTI infection, but it is not microbiologically confirmed. It may be they have other infections which are treated by the AB they consume, or that their prior AB use has supressed pathogen growth to be under the threshold for UTI identification. However, this underlines the need for greater availability of accessible, rapid diagnostic tools(42), which could prevent inappropriate AB use and help to slow growth of ABR more generally.

It is well established that excessive AB use is a core driver of ABR at individual and community scales (8), and thus, we might expect to find a correlation between longer pathways, higher levels of AB use, and higher rates of MDR in individual patients. However untangling temporality or cause and effect between AB use and MDR infections is complex, and where ABR is highly prevalent the two factors likely operate in a reinforcing cycle.

Associations between treatment complexity and MDR could be bi-directional or explained by unmeasured factors (e.g. upstream drivers/risk factors for both complex treatment seeking and ABR). In this study, the recent treatment seeking period might be a few days, or weeks, which makes it less likely that that patient’s recent AB use has directly influenced the pathogen population that is responsible for the infection, or the carriage population of the bacteria in the host that may be the source of the infection (i.e. faecal carriage to urinary tract transmission). On the other hand, the MDR infection rate is relatively high (over 50% in Tanzania and Uganda). Thus, any link we do observe between AB use and MDR UTI is more likely to be explained by primary resistance: that is, a patient acquiring an MDR UTI at the outset, and struggling to treat it, end up taking more treatment steps and more ABs. It is also worth noting that we only observe a direct association between recent AB use and MDR in Uganda, and not the other countries. However, given that the prevalence of MDR UTI in Uganda sample was high (57%) this suggests that at least, in some settings, higher MDR might be driving longer and more complex pathways, and thus higher costs and suffering for healthcare systems and patients. We recommend further research on this dynamic, that quantify the consequences of AMR for health systems and individual’s treatment seeking behaviour.

Our results also suggest the importance of considering contextual effects in unravelling the social and behavioural drivers of ABR. Country-specific analyses suggest that treatment seeking, MDR rates and the relationship between the two differ by place. Furthermore, the inclusion of variables which proxy for context (e.g. country) account for more variation in MDR than individual factors or behaviours. There are many reasons why geographic, social and cultural context should be important (14). For example, the more complex pathways we observe in Tanzania and Uganda may be a function of the shared structure of the healthcare system in those countries, or related to geographic features of the sites, which affect healthcare accessibility. The recruitment sites in Tanzania and Uganda were more rural, whereas the majority of the Kenyan samples are from the capital Nairobi. Likewise, spatial patterning of ABR is influenced by processes of bacterial evolution and transmission, affected by shared structural drivers such as sanitation (43,44). It is relevant that we do not observe consistent relationships between individual factors such as education and gender and MDR across country settings. This raises the question of whether the AMR nexus, its social patterning, and social inter-relations is contextually specific, or whether there are generalizable relationships to identify. For example, whether there are measurable area- factors which operate as upstream drivers of both treatment seeking and ABR, such as poverty(45). Future studies using HATUA data will explore these issues.

This study is novel in bridging several methodological and disciplinary divides. We bring together theoretically grounded understandings of patient behaviours in LMICs, generated largely from qualitative work (15,24), combine these with empiric diagnostic data, and analyse these using statistical and data-driven computer science techniques, typically used to derive patient flows from electronic health records in high-income countries. Thus, this is the first application of process mining to incorporate the breadth of formal and non-formal providers in LMICs. The advantage of taking a data-driven approach to summarize behavioral complexity is that it imposes minimal pre-conceived expectations, enables engaging and detailed visualizations which allow analysts and policy makers to easily identify important elements to concentrate on. We also applied sequence analysis to take account of complex sequencing or different elements of pathways and cluster patients together for further analysis. We were inevitably limited by the self-reported nature of the data and the structure of the questionnaire used to collect them. One opportunity not exploited here is the inclusion of time stamps in sequences (these were unavailable in the HATUA dataset), thus we assumed all steps were equally spaced in time, which may hide important aspects. Future studies in LMICs could try to draw on existing medical records systems where available or use digital traces to construct an understanding of systems of patient flow which take account of complexity and non-formal healthcare providers.

This study has some limitations. The linked quantitative patient sample was representative of adults attending mainly public outpatient services with UTI-like symptoms. It is likely those sampled comprised people more likely to attend clinics, thus use of other providers may be underestimated. However, patient populations are an important subgroup for possible interventions, and UTI is both prevalent and commonly treated with ABs in this region. Other data collected by the HATUA Consortium used community focus groups, and many of the same themes -–of repeat clinic visits and challenges in obtaining appropriate medications– emerged there(12). There were also proportionally more respondents from urban higher-level referral facilities in Kenya compared to the other countries. Finally, we analysed a simple set of individual level predictors, with a focus on behavioural antecedents of clinic attendance, but our own qualitative work (12,13) suggests that a richer set of covariates at individual, household and community are necessary to understand the burden of ABR, as well as longitudinal follow-up. This will be addressed in future HATUA studies which take account of the inter-related and multi-scalar nature of ABR drivers.

## Conclusion

This novel interdisciplinary mixed-methods study combining social science and microbiological data highlights the complexities of treatment-seeking for UTI care in East Africa, and the potential for higher rates of ABR to complicate patient treatment seeking. In Tanzania and Uganda, where more than half of UTI patients surveyed have MDR UTIs and overall prevalence of ABR is high(20), repeated treatment failures involving a plethora of providers are an inevitable consequence of patients seeking to alleviate their symptoms.

Such frustrating and expensive cycles of treatment place a heavy burden on already struggling individuals and healthcare structures, and feed into a vicious cycle of ineffective AB use and further selection pressure for ABR in pathogen populations. Solutions for arresting this cycle are likely multifaceted, including improving access to rapid diagnostic and susceptibility testing for UTIs, appropriate AB treatment, effective AB stewardship, and addressing the upstream drivers of infection, through behavioural factors like hygiene and environmental determinants such as sanitation(43).

## Data Availability

All data produced in the present work are contained in the manuscript.

## Acknowledgements

HATUA consortium includes all named authors plus: Elliott, Alison; Lynch; Andy; Stelling, John; Maina, John; Kansiime; Catherine; Aduda, Annette

## Ethics approval and consent to participate

The study received ethical approval from the University of St Andrews, UK (number MD14548, 10/09/19); National Institute for Medical Research, Tanzania (number 2831, updated 26/07/19); CUHAS/BMC Research Ethics and Review Committee (number CREC /266/2018, updated on 02/2019); Mbeya Medical Research and Ethics Committee (number SZEC-2439/R.A/V.1/303030); Kilimanjaro Christian Medical College, Tanzania (number 2293, updated 14/08/19); Uganda National Council for Science and Technology (number HS2406, 18/06/18); Makerere University, Uganda (number 514, 25/04/18); and Kenya Medical Research Institute (04/06/19, Scientific and Ethics Review Committee (SERU) number KEMRI/SERU/CMR/P00112/3865 V.1.2). For Uganda, administrative letters of support were obtained from the district health officers to allow the research to be conducted in the respective hospitals and health centres. All participants provided written informed consent to participate.

## Availability of data and materials

The datasets generated and/or analysed during the current study are not publicly available due ethical and data access agreements with individual country ethical boards but are available from the corresponding author on reasonable request.

## Author contributions

KS: Conceptualisation, data curation, formal analysis, investigation, visualisation, writing- original draft preparation

KK: conceputalisation, data curation, funding acquisition, methodology, supervision, writing- original draft preparation, review and editing

AM: conceptualisation, methodology, supervision, writing- review and editing

MK: conceptualisation, funding acquisition, supervision, writing- review and editing MFM: data curation, methodology, resources, supervision, writing- review and editing

SEM data curation, funding acquisition, methodology, resources, supervision, writing- review and editing

JM data curation, funding acquisition, methodology, resources, supervision, writing- review and editing

SN data curation, funding acquisition, methodology, resources, supervision, writing- review and editing

BA data curation, funding acquisition, methodology, resources, supervision, writing- review and editing

JB data curation, methodology, resources, supervision, writing- review and editing JK data curation, methodology, resources, supervision, writing- review and editing DLG data curation, writing- review and editing

XK data curation, writing- review and editing AMB data curation, writing- review and editing MAAA data curation, writing- review and editing KF review and editing

SHG funding acquisition, methodology, resources, writing- review and editing

BTM data curation, funding acquisition, methodology, resources, supervision, writing- review and editing

WS funding acquisition, writing- review and editing GK funding acquisition, writing- review and editing DA funding acquisition, writing- review and editing VAS funding acquisition, writing- review and editing AS Project administration, writing- review and editing

DJS funding acquisition, supervision, writing- review and editing

MTGH conceptualisation, methodology, resources, funding acquisition, supervision, writing- review and editing

## References

1. Holmes AH, Moore LSP, Sundsfjord A, Steinbakk M, Regmi S, Karkey A, et al. Understanding the mechanisms and drivers of antimicrobial resistance. The Lancet. 2016 Jan 9;387(10014):176–87.

2. Murray CJ, Ikuta KS, Sharara F, Swetschinski L, Robles Aguilar G, Gray A, et al. Global burden of bacterial antimicrobial resistance in 2019: a systematic analysis. The Lancet. 2022 Feb 12;399(10325):629–55.

3. O’Neill J. Antimicrobial Resistance : Tackling a crisis for the health and wealth of [] nations. 2014.

4. D’Costa VM, King CE, Kalan L, Morar M, Sung WWL, Schwarz C, et al. Antibiotic resistance is ancient. Nature. 2011 Sep;477(7365):457–61.

5. Davies J, Davies D. Origins and Evolution of Antibiotic Resistance. Microbiol Mol Biol Rev. 2010 Sep;74(3):417–33.

6. Goossens H, Ferech M, Vander Stichele R, Elseviers M. Outpatient antibiotic use in Europe and association with resistance: a cross-national database study. The Lancet. 2005 Feb 12;365(9459):579–87.

7. Austin DJ, Kristinsson KG, Anderson RM. The relationship between the volume of antimicrobial consumption in human communities and the frequency of resistance. Proc Natl Acad Sci. 1999 Feb 2;96(3):1152–6.

8. Bell BG, Schellevis F, Stobberingh E, Goossens H, Pringle M. A systematic review and meta-analysis of the effects of antibiotic consumption on antibiotic resistance. BMC Infect Dis. 2014 Jan 9;14(1):13.

9. Chatterjee A, Modarai M, Naylor NR, Boyd SE, Atun R, Barlow J, et al. Quantifying drivers of antibiotic resistance in humans: a systematic review. Lancet Infect Dis. 2018;18(12):e368–78.

10. Charani E, McKee M, Ahmad R, Balasegaram M, Bonaconsa C, Merrett GB, et al. Optimising antimicrobial use in humans – review of current evidence and an interdisciplinary consensus on key priorities for research. Lancet Reg Health - Eur. 2021 Aug 1;7:100161.

11. Muela SH, Ribera JM, Mushi AK, Tanner M. Medical syncretism with reference to malaria in a Tanzanian community. Soc Sci Med 1982. 2002 Aug;55(3):403–13.

12. Green DL, Keenan K, Fredricks KJ, Huque SI, Mushi MF, Kansiime C, et al. The role of multidimensional poverty in antibiotic misuse: a mixed-methods study of self-medication and non-adherence in Kenya, Tanzania, and Uganda. Lancet Glob Health. 2023 Jan 1;11(1):e59–68.

13. Keenan K, Fredricks KJ, Ahad MAA, Neema S, Mwanga JR, Kesby M, et al. Unravelling patient pathways in the context of antibacterial resistance in East Africa [Internet]. In Review; 2022 Aug [cited 2022 Sep 9]. Available from: https://www.researchsquare.com/article/rs-1933132/v1

14. Do NTT, Vu HTL, Nguyen CTK, Punpuing S, Khan WA, Gyapong M, et al. Community-based antibiotic access and use in six low-income and middle-income countries: a mixed-method approach. Lancet Glob Health [Internet]. 2021 Mar 10 [cited 2021 Mar 18]; Available from: https://linkinghub.elsevier.com/retrieve/pii/S2214109X21000243

15. Lucas PJ, Uddin MR, Khisa N, Salim Akter SM, Unicomb L, Nahar P, et al. Pathways to antibiotics in Bangladesh: A qualitative study investigating how and when households access medicine including antibiotics for humans or animals when they are ill. PLoS ONE. 2019 Nov 1;14(11):e0225270.

16. Torres NF, Chibi B, Kuupiel D, Solomon VP, Mashamba-Thompson TP, Middleton LE. The use of non-prescribed antibiotics; prevalence estimates in low-and-middle-income countries. A systematic review and meta-analysis. Arch Public Health. 2021 Dec 1;79(1):1–15.

17. Ocan M, Obuku EA, Bwanga F, Akena D, Richard S, Ogwal-Okeng J, et al. Household antimicrobial self-medication: A systematic review and meta-analysis of the burden, risk factors and outcomes in developing countries. BMC Public Health. 2015 Aug 1;15(1):1– 11.

18. Aslam A, Gajdács M, Zin CS, Rahman NSA, Ahmed SI, Zafar MZ, et al. Evidence of the practice of self-medication with antibiotics among the lay public in low-and middle-income countries: A scoping review. Antibiotics. 2020 Sep 1;9(9):1–17.

19. Global access to antibiotics without prescription in community pharmacies: A systematic review and meta-analysis - ScienceDirect [Internet]. [cited 2022 Sep 8]. Available from: https://www.sciencedirect.com/science/article/pii/S0163445318302123?casa_token=f-bOkqgbJZ4AAAAA:yorgRk2UGGmcuMS5EfIzVrX6Tvv3TRWg16Sgl-POqc47eoW4ebUdeoFUUFInAe4G0C1zPDO0RQ

20. Leopold SJ, van Leth F, Tarekegn H, Schultsz C. Antimicrobial drug resistance among clinically relevant bacterial isolates in sub-Saharan Africa: A systematic review. J Antimicrob Chemother. 2014 Sep 1;69(9):2337–53.

21. Ndaki PM, Mushi MF, Mwanga JR, Konje ET, Ntinginya NE, Mmbaga BT, et al. Dispensing Antibiotics without Prescription at Community Pharmacies and Accredited Drug Dispensing Outlets in Tanzania: A Cross-Sectional Study. Antibiot 2021 Vol 10 Page 1025. 2021 Aug 23;10(8):1025.

22. Mburu CM, Bukachi SA, Shilabukha K, Tokpa KH, Ezekiel M, Fokou G, et al. Determinants of treatment-seeking behavior during self-reported febrile illness episodes using the socio-ecological model in Kilombero District, Tanzania. BMC Public Health. 2021 Dec 1;21(1):1–11.

23. Haenssgen MJ, Ariana P. Healthcare access: A sequence-sensitive approach. SSM - Popul Health. 2017 Dec 1;3:37–47.

24. Hanson CL, Osberg M, Brown J, Durham G, Chin DP. Conducting Patient-Pathway Analysis to Inform Programming of Tuberculosis Services: Methods. J Infect Dis. 2017 Nov 6;216(suppl_7):S679–85.

25. Coast E, Norris AH, Moore AM, Freeman E. Trajectories of women’s abortion-related care: A conceptual framework. Soc Sci Med. 2018 Mar 1;200:199–210.

26. Zøylner IA, Lomborg K, Christiansen PM, Kirkegaard P. Surgical breast cancer patient pathway: Experiences of patients and relatives and their unmet needs. Health Expect. 2019 Apr 1;22(2):262–72.

27. Asiimwe BB, Kiiru J, Mshana SE, Neema S, Keenan K, Kesby M, et al. Protocol for an interdisciplinary cross-sectional study investigating the social, biological and community-level drivers of antimicrobial resistance (AMR): Holistic Approach to Unravel Antibacterial Resistance in East Africa (HATUA). BMJ Open. 2021 Mar;11(3):e041418.

28. Izett-Kay M, Barker KL, McNiven A, Toye F. Experiences of urinary tract infection: A systematic review and meta-ethnography. Neurourol Urodyn [Internet]. 2022 [cited 2022 Mar 23]; Available from: https://onlinelibrary.wiley.com/doi/full/10.1002/nau.24884

29. Ozturk R, Murt A. Epidemiology of urological infections: a global burden. World J Urol. 2020;38(11):2669–79.

30. Rojas E, Munoz-Gama J, Sepúlveda M, Capurro D. Process mining in healthcare: A literature review. J Biomed Inform. 2016 Jun 1;61:224–36.

31. Magiorakos AP, Srinivasan A, Carey RB, Carmeli Y, Falagas ME, Giske CG, et al. Multidrug-resistant, extensively drug-resistant and pandrug-resistant bacteria: an international expert proposal for interim standard definitions for acquired resistance. Clin Microbiol Infect. 2012 Mar 1;18(3):268–81.

32. Dixon J, MacPherson E, Manyau S, Nayiga S, Khine Zaw Y, Kayendeke M, et al. The ‘Drug Bag’ method: lessons from anthropological studies of antibiotic use in Africa and South- East Asia. Glob Health Action. 2019 Dec;12(sup1):1639388.

33. Aassve Francesco C Billari AE Raffaella Piccarreta AA, Aassve ISER A, Billari FC, Piccarreta IMQ R. Strings of Adulthood: A Sequence Analysis of Young British Women’s Work-Family Trajectories Parcours de la vie adulte : Une analyse par séquence des trajectoires travail-famille des jeunes femmes britanniques. Eur J Popul. 2007;23:369–88.

34. Cezard G, Sullivan F, Keenan K. Understanding multimorbidity trajectories in Scotland using sequence analysis | Scientific Reports. Sci Rep [Internet]. 2022 Nov 1 [cited 2023 Jan 25];12(16485). Available from: https://www.nature.com/articles/s41598-022-20546-4

35. Berti, A., van Zelst, S.J, van der Aalst, W.M.P. Process Mining for Python (PM4Py): Bridging the Gap Between Process-and Data Science. In: Proceedings of the ICPM Demo Track 2019 [Internet]. Aachen, Germany; 2019. Available from: http://ceur-ws.org/Vol-2374/

36. Verbeek HMW, Buijs JCAM, van Dongen BF, van der Aalst WMP. XES, XESame, and ProM 6. In: Soffer P, Proper E, editors. Information Systems Evolution. Berlin, Heidelberg: Springer; 2011. p. 60–75. (Lecture Notes in Business Information Processing).

37. Studer M. WeightedCluster Library Manual: A practical guide to creating typologies of trajectories in the social sciences with R. 2013 [cited 2022 Sep 6]; Available from: https://www.centre-lives.ch/fr/bibcite/reference/84

38. Cornwell B. Social Sequence Analysis: Methods and Applications. Cambridge University Press; 2015. 339 p.

39. Sundararajan R, Mwanga-Amumpaire J, King R, Ware NC. Conceptual model for pluralistic healthcare behaviour: results from a qualitative study in southwestern Uganda. BMJ Open. 2020 Apr 1;10(4):e033410.

40. Towns AM, Eyi SM, Andel T van. Traditional Medicine and Childcare in Western Africa: Mothers’ Knowledge, Folk Illnesses, and Patterns of Healthcare-Seeking Behavior. PLOS ONE. 2014 Aug 22;9(8):e105972.

41. Moshabela M, Bukenya D, Darong G, Wamoyi J, McLean E, Skovdal M, et al. Traditional healers, faith healers and medical practitioners: the contribution of medical pluralism to bottlenecks along the cascade of care for HIV/AIDS in Eastern and Southern Africa. Sex Transm Infect [Internet]. 2017 Jul 1 [cited 2023 Jan 25];93(Suppl 3). Available from: https://sti.bmj.com/content/93/Suppl_3/e052974

42. Fleming KA, Horton S, Wilson ML, Atun R, DeStigter K, Flanigan J, et al. The Lancet Commission on diagnostics: transforming access to diagnostics. The Lancet. 2021 Nov 27;398(10315):1997–2050.

43. Fletcher S. Understanding the contribution of environmental factors in the spread of antimicrobial resistance. Environ Health Prev Med. 2015 Jul 1;20(4):243–52.

44. Cocker D, Chidziwisano K, Mphasa M, Mwapasa T, Lewis JM, Rowlingson B, et al. Investigating risks for human colonisation with extended spectrum beta-lactamase producing E. coli and K. pneumoniae in Malawian households: a one health longitudinal cohort study [Internet]. medRxiv; 2022 [cited 2022 Nov 22]. p. 2022.08.16.22278508. Available from: https://www.medrxiv.org/content/10.1101/2022.08.16.22278508v1

45. Okeke IN. Poverty and root causes of resistance in developing countries. In: Antimicrobial Resistance in Developing Countries [Internet]. Springer New York; 2010 [cited 2021 Mar 18]. p. 27–35. Available from: https://link.springer.com/chapter/10.1007/978-0-387-89370-9_3

