## Supplementary material for "Treatment seeking behaviours, antibiotic use and relationships to multi-drug resistance: A study of urinary tract infection patients in Kenya, Tanzania and Uganda"

**Table S1.** Patient recruitment sites in Kenya, Tanzania, and Uganda.

| **Country/site** | **Number of facilities** | **Source of funding** | **Levels recruited from^1^** |
| --- | --- | --- | --- |
| **Kenya** |  |  |  |
| Makueni | 1 | Public | 5 |
| Nairobi | 4 | Public and private | 3-5, National |
| Nanyuki | 1 | Public | 4 |
| **Tanzania** |  |  |  |
| Kilimanjaro/Moshi | 3 | Public and private | 2,3, and 5 |
| Mbeya | 2 | Public and private | 3 and 4 |
| Mwanza | 5 | Public and private | 2,3, and 5 |
| **Uganda** |  |  |  |
| Mbarara | 3 | Public | 3 and 5 |
| Nakapiripirit | 3 | Public | 2 and 3 |
| Nakasongola | 3 | Public and private | 3 and 4 |

^1^ Levels of facilities are identified in each country following the Kenya Health Policy 2014-2013 (<http://publications.universalhealth2030.org/uploads/kenya_health_policy_2014_to_2030.pdf>); Tanzanian Health Sector Strategic Plan, <https://p4h.world/en/node/11813#:~:text=2026%20%7C%20P4H%20Network-,Tanzania%20fifth%20health%20sector%20strategic%20plan%20(HSSPV)%2D%202021%2D2026,coverage%20(UHC)%20by%202030>

and the Ugandan Hospital and Health Centre IV census survey (<http://library.health.go.ug/publications/health-infrastructure/uganda-hospital-and-health-centre-iv-census-survey>)

**Table S2.** Antibiotics considered for MDR calculations.

| **Gram negative** | **Amoxicillin/**  **clavulanate**  **(AMC)** | **Ampicillin**  **(AMP)** | **Ceftazidime/**  **Ceftriaxone**  **(CAZ/CRO)** | **Ciprofloxacin**  **(CIP)** | **Gentamicin**  **(GEN)** | **Nitrofurantoin**  **(NIT)** | **Trimethoprim**  **(TMP)** |  | |
| --- | --- | --- | --- | --- | --- | --- | --- | --- | --- |
| ***E. coli*** | AMC | AMP | CAZ/CRO | CIP | GEN | NIT | TMP |  | |
| ***Shigella* spp.** | AMC | AMP | CAZ/CRO | CIP | GEN | NIT | TMP |  | |
| ***Proteus* spp.** | AMC | AMP | CAZ/CRO | CIP | GEN | NIT | TMP |  | |
| ***Salmonella* spp.** | AMC | AMP | CAZ/CRO | CIP | GEN | NIT | TMP |  | |
| ***Serratia* spp.** | - | - | CAZ/CRO | CIP | GEN | NIT | TMP |  | |
| ***Klebsiella* spp.** | AMC | - | CAZ/CRO | CIP | GEN | NIT | TMP |  | |
| ***Citrobacter* spp.** | **-** | - | CAZ/CRO | CIP | GEN | NIT | TMP |  | |
| ***Enterobacter* spp.** | - | - | CAZ/CRO | CIP | GEN | NIT | TMP |  | |
| ***Morganella* spp.** | **-** | **-** | CAZ/CRO | CIP | GEN | NIT | TMP |  | |
| ***Pantoea* spp.** | - | - | CAZ/CRO | CIP | GEN | NIT | TMP |  | |
| ***Providencia* spp.** | - | - | CAZ/CRO | CIP | GEN | NIT | TMP |  | |
| ***Acinetobacter* spp.** | - | - | CAZ/CRO | CIP | GEN | - | TMP |  | |
| ***Pseudomonas* spp.** | - | - | CAZ | CIP | GEN | - | - |  | |
| **Gram positive** | **Cefoxitin**  **(FOX)** | **Erythromycin**  **(ERY)** | **Linezolid**  **(LNZ)** | **Ciprofloxacin**  **(CIP)** | **Gentamicin**  **(GEN)** | **Nitrofurantoin**  **(NIT)** | **Trimethoprim**  **(TMP)** | **Tetracycline**  **(TCY)** | **Vancomycin**  **(VAN)** |
| ***Staphylococcus* spp.** | FOX | ERY | - | CIP | GEN | NIT | TMP | TCY | - |
| ***Enterococcus* spp.** | - | ERY | LNZ | CIP | - | NIT | - | TCY | VAN |
| ***Streptococcus* spp.** | **-** | ERY | LNZ | - | - | NIT | - | TCY | VAN |

**Table S3. List of antibiotics mentioned by patients and whether they are recommended for use for treating UTI according to country National Treatment Guidelines (NTG)**

|  | **Kenya NTG^1^**  (adult outpatients- not admitted to hospital) | **Tanzania NTG^2^**  (adult outpatients- not admitted to hospital) | **Uganda NTG^3^**  (adult outpatients- not admitted to hospital) |
| --- | --- | --- | --- |
| Amoxicillin | a, b |  | c |
| Amoxicillin/ Clavulanic Acid (Amoxiclav) |  | b, c |  |
| Ampicillin | a |  | b, c |
| Ampicillin-Cloxacillin (Ampiclox) |  |  |  |
| Azithromycin (Azuma) |  |  |  |
| Ceftriaxone |  |  | b, c |
| Cefuroxime |  |  |  |
| Cephalexine |  |  |  |
| Ciprofloxacin |  | a, b | a, b |
| Cotrimoxozole (Septrine) | a, b |  |  |
| Doxycycline |  |  |  |
| Erythromycin |  |  |  |
| Gentamicin |  |  | b, c |
| Levofloxacin |  |  |  |
| Metronidazole (Flagyl) |  |  |  |
| Nitrofurantoin | a |  | a, c |
| Tetracycline |  |  |  |
| Tinidazole |  |  |  |

a= uncomplicated lower UTI

b= uncomplicated upper UTI (includes for Uganda, first and second line options)

c= treatments recommended for pregnant women only

**^1^** [**^http://guidelines.health.go.ke:8000/media/Clinical_Guidelines_Vol_II_Final.pdf^**](http://guidelines.health.go.ke:8000/media/Clinical_Guidelines_Vol_II_Final.pdf)

**^2^** [**^https://hssrc.tamisemi.go.tz/storage/app/uploads/public/5ab/e9b/b21/5abe9bb216267130384889.pdf^**](https://hssrc.tamisemi.go.tz/storage/app/uploads/public/5ab/e9b/b21/5abe9bb216267130384889.pdf)

**^3^** [**^https://www.prb.org/wp-content/uploads/2018/05/Uganda-Clinical-Guidelines-2016-National-Guidelines-for-Management-of-Common-Conditions.pdf^**](https://www.prb.org/wp-content/uploads/2018/05/Uganda-Clinical-Guidelines-2016-National-Guidelines-for-Management-of-Common-Conditions.pdf)

**Table S4: Characteristics of the patient sample used for the two stages of the analysis**

|  |  | **Analysis sample: Pathway characteristics** | | **Analysis sample: Associations with MDR** | |
| --- | --- | --- | --- | --- | --- |
|  |  | **N** | **%** | **N** | **%** |
| **Country** | Kenya | 1,591 | 24.9 | 835 | 42.9 |
|  | Tanzania | 3,046 | 47.7 | 672 | 34.5 |
|  | Uganda | 1,751 | 27.4 | 439 | 22.6 |
| **Age** | <25 | 1,695 | 26.5 | 526 | 27.0 |
|  | 25-34 | 2,114 | 33.1 | 673 | 34.6 |
|  | 35-44 | 1,021 | 16.0 | 175 | 9.0 |
|  | 45-54 | 632 | 9.9 | 89 | 4.6 |
|  | 55-64 | 373 | 5.8 | 289 | 14.9 |
|  | 65+ | 553 | 8.7 | 194 | 10.0 |
| **Gender** | Male | 1,367 | 21.4 | 277 | 14.2 |
|  | Female | 5,021 | 78.6 | 1,669 | 85.8 |
| **Education** | None | 965 | 15.1 | 282 | 14.5 |
|  | Primary | 2,525 | 39.5 | 658 | 33.8 |
|  | Secondary | 1,984 | 31.1 | 664 | 34.1 |
|  | Higher | 914 | 14.3 | 342 | 17.6 |
| **Treatment steps** | 1(straight to clinic) | 2,879 | 45.1 | 985 | 50.6 |
|  | 2 | 1,835 | 28.7 | 558 | 28.7 |
|  | 3+ | 1,674 | 26.2 | 403 | 20.7 |
| **AB use in pathway** | No | 4,505 | 70.5 | 1,444 | 74.2 |
|  | Yes | 1,883 | 29.5 | 502 | 25.8 |
| **AB use past 6m** | No | 2,482 | 38.9 | 804 | 41.3 |
|  | Yes | 3,906 | 61.1 | 1,142 | 58.7 |
| **UTI status** | Negative | 4,194 | 65.7 | 0 | 0 |
|  | Positive | 2,194 | 34.3 | 1,946 | 100 |
| **MDR status** | Negative |  |  | 1,014 | 52.1 |
|  | Positive |  |  | 932 | 47.9 |
| **TOTAL** |  | 6,388 | 100.0 | 1,946 | 100.0 |

**Table S5: Kenya: characteristics of the patient sample used for the two stages of the analysis**

|  |  | **Analysis sample: Pathway characteristics** | | **Analysis sample: Associations with MDR** | |
| --- | --- | --- | --- | --- | --- |
|  |  | **N** | **%** | **N** | **%** |
| **Age** | <25 | 433 | 27.2 | 245 | 29.3 |
|  | 25-34 | 753 | 47.3 | 389 | 46.6 |
|  | 35-44 | 254 | 16.0 | 39 | 4.7 |
|  | 45-54 | 85 | 5.3 | 11 | 1.3 |
|  | 55-64 | 25 | 1.6 | 123 | 14.7 |
|  | 65+ | 41 | 2.6 | 28 | 3.4 |
| **Gender** | Male | 258 | 16.2 | 66 | 7.9 |
|  | Female | 1,333 | 83.8 | 769 | 92.1 |
| **Education** | None | 19 | 1.2 | 11 | 1.3 |
|  | Primary | 195 | 12.3 | 99 | 11.9 |
|  | Secondary | 889 | 55.9 | 460 | 55.1 |
|  | Higher | 488 | 30.7 | 265 | 31.7 |
| **Treatment steps** | 1(straight to clinic) | 1,041 | 65.4 | 574 | 68.7 |
|  | 2 | 450 | 28.3 | 224 | 26.8 |
|  | 3+ | 100 | 6.3 | 37 | 4.4 |
| **AB use in pathway** | No | 1,303 | 81.9 | 690 | 82.6 |
|  | Yes | 288 | 18.1 | 145 | 17.4 |
| **AB use past 6m** | No | 758 | 47.6 | 370 | 44.3 |
|  | Yes | 833 | 52.4 | 465 | 55.7 |
| **UTI status** | Negative | 716 | 45.0 | 0 | 0 |
|  | Positive | 875 | 55.0 | 835 | 100 |
| **MDR status** | Negative |  |  | 553 | 66.2 |
|  | Positive |  |  | 282 | 33.8 |
| **TOTAL** |  | 1,591 | 100.0 | 835 | 100.0 |

**Table S6: Tanzania: characteristics of the patient sample used for the two stages of the analysis**

|  |  | **Analysis sample: Pathway characteristics** | | **Analysis sample: Associations with MDR** | |
| --- | --- | --- | --- | --- | --- |
|  |  | **N** | **%** | **N** | **%** |
| **Age** | <25 | 675 | 22.2 | 139 | 20.7 |
|  | 25-34 | 799 | 26.2 | 140 | 20.8 |
|  | 35-44 | 469 | 15.4 | 101 | 15.0 |
|  | 45-54 | 375 | 12.3 | 78 | 11.6 |
|  | 55-64 | 278 | 9.1 | 65 | 9.7 |
|  | 65+ | 450 | 14.8 | 149 | 22.2 |
| **Gender** | Male | 832 | 27.3 | 158 | 23.5 |
|  | Female | 2,214 | 72.7 | 514 | 76.5 |
| **Education** | None | 349 | 11.5 | 113 | 16.8 |
|  | Primary | 1,678 | 55.1 | 400 | 59.5 |
|  | Secondary | 758 | 24.9 | 122 | 18.2 |
|  | Higher | 261 | 8.6 | 37 | 5.5 |
| **Treatment steps** | 1(straight to clinic) | 1,051 | 34.5 | 221 | 32.9 |
|  | 2 | 795 | 26.1 | 183 | 27.2 |
|  | 3+ | 1,200 | 39.4 | 268 | 39.9 |
| **AB use in pathway** | No | 2,056 | 67.5 | 467 | 69.5 |
|  | Yes | 990 | 32.5 | 205 | 30.5 |
| **AB use past 6m** | No | 510 | 16.7 | 139 | 20.7 |
|  | Yes | 2,536 | 83.3 | 533 | 79.3 |
| **UTI status** | Negative | 2,214 | 72.7 | 0 | 0 |
|  | Positive | 832 | 27.3 | 672 | 100 |
| **MDR status** | Negative |  |  | 272 | 40.5 |
|  | Positive |  |  | 400 | 59.5 |
| **TOTAL** |  | 3,046 | 100.0 | 672 | 100.0 |

**Table S7: Uganda: characteristics of the patient sample used for the two stages of the analysis**

|  |  | **Analysis sample: Pathway characteristics** | | **Analysis sample: Associations with MDR** | |
| --- | --- | --- | --- | --- | --- |
|  |  | **N** | **%** | **N** | **%** |
| **Age** | <25 | 587 | 33.5 | 142 | 32.3 |
|  | 25-34 | 562 | 32.1 | 144 | 32.8 |
|  | 35-44 | 298 | 17.0 | 65 | 14.8 |
|  | 45-54 | 172 | 9.8 | 58 | 13.2 |
|  | 55-64 | 70 | 4.0 | 13 | 3.0 |
|  | 65+ | 62 | 3.5 | 17 | 3.9 |
| **Gender** | Male | 277 | 15.8 | 53 | 12.1 |
|  | Female | 1,474 | 84.2 | 386 | 87.9 |
| **Education** | None | 597 | 34.1 | 158 | 36.0 |
|  | Primary | 652 | 37.2 | 159 | 36.2 |
|  | Secondary | 337 | 19.2 | 82 | 18.7 |
|  | Higher | 165 | 9.4 | 40 | 9.1 |
| **Treatment steps** | 1(straight to clinic) | 787 | 44.9 | 190 | 43.3 |
|  | 2 | 590 | 33.7 | 151 | 34.4 |
|  | 3+ | 374 | 21.4 | 98 | 22.3 |
| **AB use in pathway** | No | 1,146 | 65.4 | 287 | 65.4 |
|  | Yes | 605 | 34.6 | 152 | 34.6 |
| **AB use past 6m** | No | 1,214 | 69.3 | 295 | 67.2 |
|  | Yes | 537 | 30.7 | 144 | 32.8 |
| **UTI status** | Negative | 1,264 | 72.2 | 0 | 0 |
|  | Positive | 487 | 27.8 | 439 | 100 |
| **MDR status** | Negative |  |  | 189 | 43.1 |
|  | Positive |  |  | 250 | 56.9 |
| **TOTAL** |  | 1,751 | 100.0 | 439 | 100.0 |

**Figure S1. Clustering quality for subgroup of sequences including 2 steps**

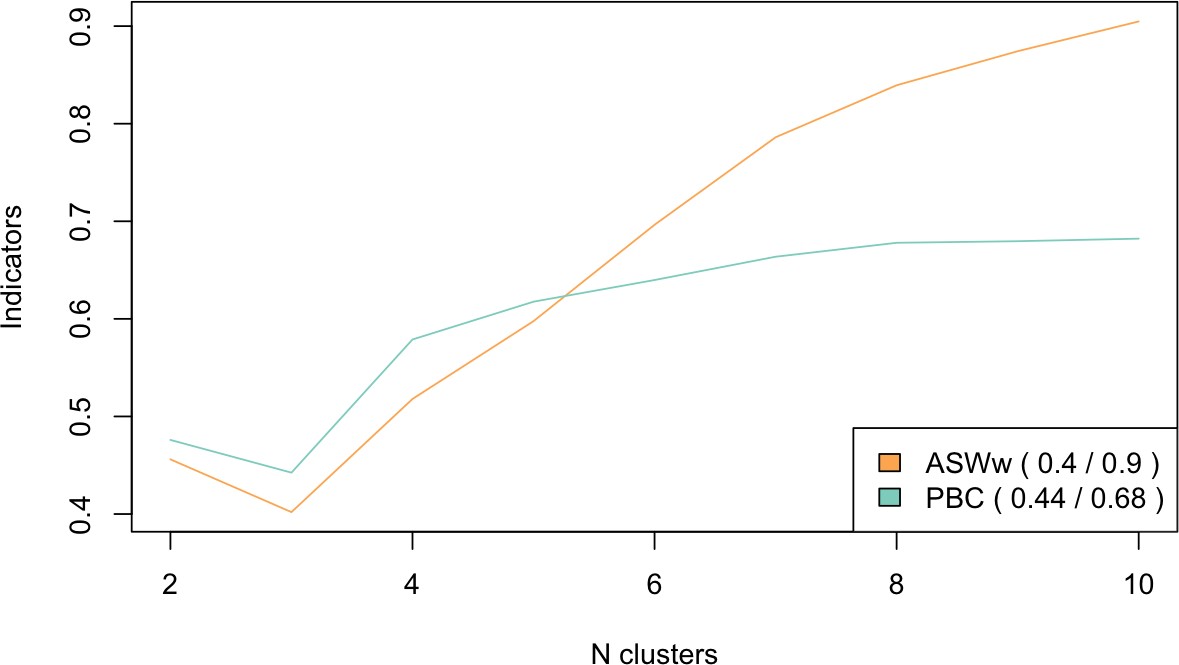

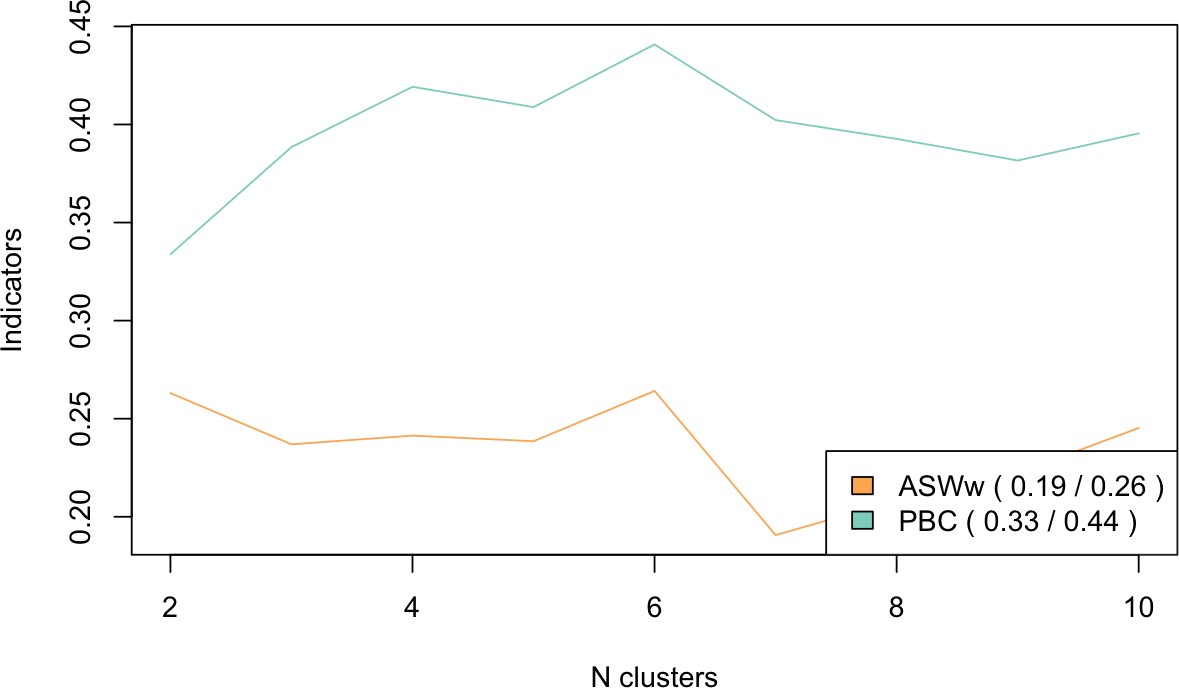
**Figure S2. Clustering quality for subgroup of sequences including 3 or more steps**

**Figure S3. Characteristics of the 10 clusters detected by sequence analysis.**

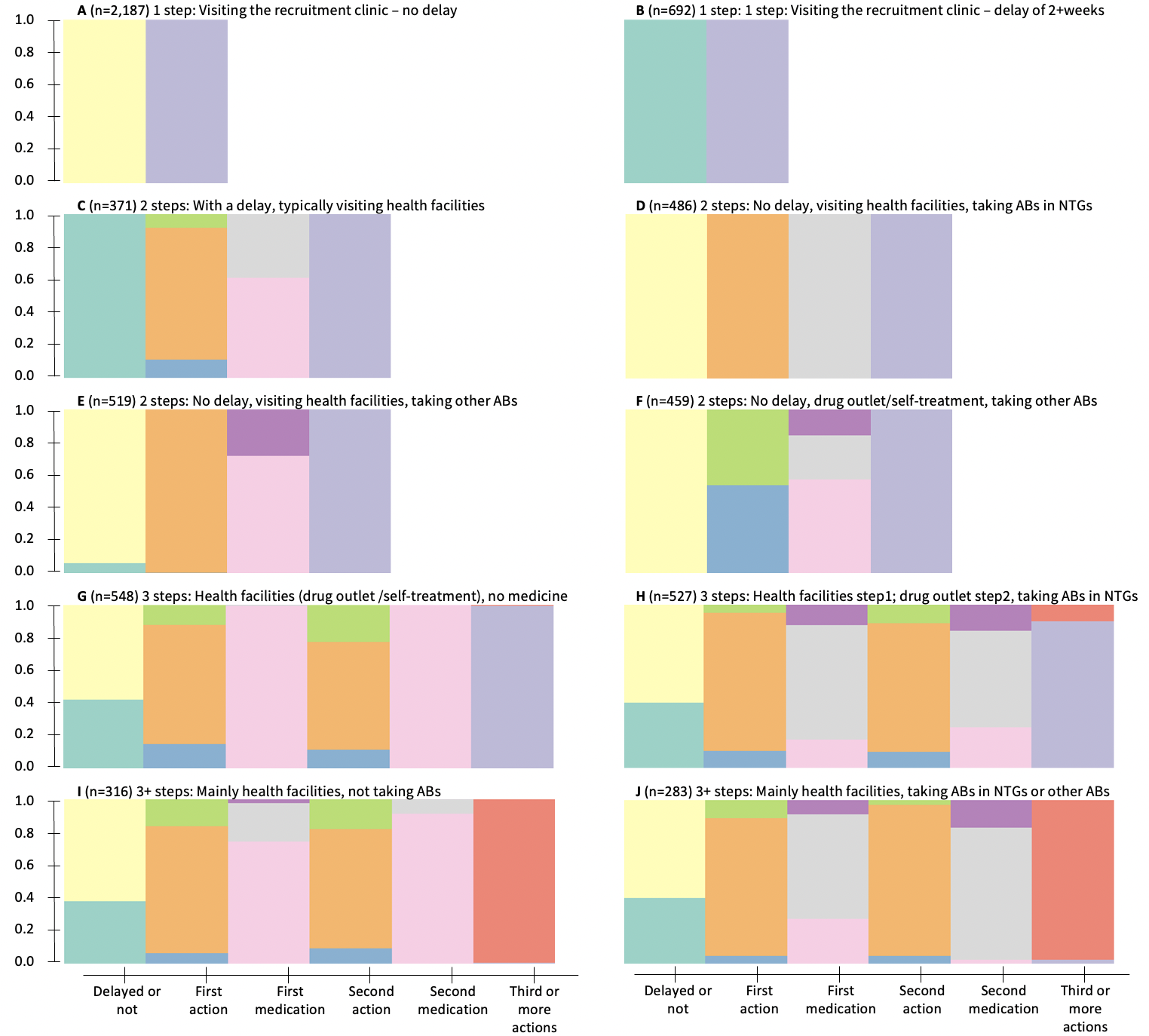

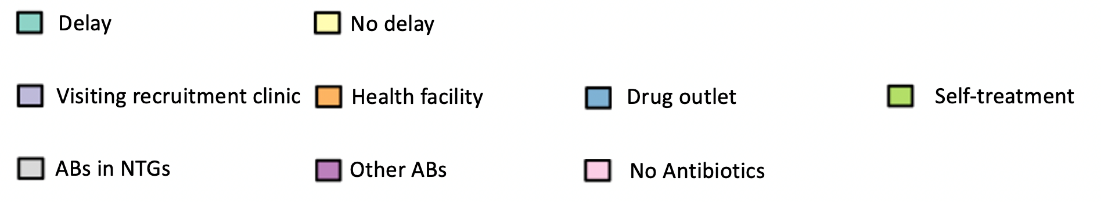

Notes: The x axis represents pathway steps and characteristics, and the y axis shows the proportion of individuals in the cluster with that pathway characteristic. For example, all patients in cluster D had no delay, went to healthcare facilities as their first step, and got ABs from NTGs, before visiting the recruitment clinics. Subjects included in cluster F also had no delay, but for the first step over half visited pharmacies/drug shops, and some of those obtained ABs, or no ABs, before showing up at the recruitment clinic.

**Table S8. Characteristics of patients included and excluded from the analysis samples**

|  |  | **Analysis sample 1** | | **Analysis sample 2 – UTI + and MDR data** | |
| --- | --- | --- | --- | --- | --- |
|  |  | **Included** | **Excluded (N%)** | **Included** | **Excluded N(%)** |
| **Country** | **Kenya** | 1,591 | 70(4.2) | 835 | 826(49.7) |
|  | **Tanzania** | 3,046 | 144(4.5) | 672 | 2518(78.9) |
|  | **Uganda** | 1,751 | 6(0.3) | 439 | 1318(75) |
| **Age** | **<25** | 1,695 | 47(2.7) | 526 | 1216(69.8) |
|  | **25-34** | 2,114 | 72(3.3) | 673 | 1513(69.2) |
|  | **35-44** | 1,021 | 27(2.6) | 289 | 759(72.4) |
|  | **45-54** | 632 | 9(1.4) | 175 | 466(72.7) |
|  | **55-64** | 373 | 5(1.3) | 89 | 289(76.5) |
|  | **65+** | 553 | 15(2.6) | 194 | 374(65.8) |
| **Gender** | **Male** | 1,367 | 42(3) | 277 | 1132(80.3) |
|  | **Female** | 5,021 | 178(3.4) | 1669 | 3530(67.9) |
| **Education** | **None** | 965 | 33(3.3) | 282 | 716(71.7) |
|  | **Primary** | 2,525 | 103(3.9) | 658 | 1970(75) |
|  | **Secondary** | 1,984 | 50(2.5) | 664 | 1370(67.4) |
|  | **Higher** | 914 | 29(3.1) | 342 | 601(63.7) |
| **TOTAL** |  | 6,388 | 220 (3.3) | 1,946 | 4,662 (70.1) |

**Table S9: Bivariate associations between patient characteristics and MDR, stratified by country**

|  |  |  | MDR-  n(%) | MDR+  n(%) | $\chi^{2}$, p value |
| --- | --- | --- | --- | --- | --- |
| **Gender** | Kenya | Male | 40 (60.6) | 26 (39.4) | 0.8 (0.384) |
|  |  | Female | 513 (66.7) | 256 (33.3) |  |
|  | Tanzania | Male | 49 (31.0) | 109 (69.0) | 7.2 (0.007) |
|  |  | Female | 223 (43.4) | 291 (56.6) |  |
|  | Uganda | Male | 26 (49.1) | 27 (50.9) | 0.6 (0.427) |
|  |  | Female | 163 (42.2) | 223 (57.8) |  |
| **Age** | Kenya | <25 | 172 (70.2) | 73 (29.8) | 6.4 (0.271) |
|  |  | 25-34 | 256 (65.8) | 133 (34.2) |  |
|  |  | 35-44 | 81 (65.9) | 42 (34.1) |  |
|  |  | 45-54 | 20 (51.3) | 19 (48.7) |  |
|  |  | 55-64 | 6 (54.5) | 5 (45.5) |  |
|  |  | 65+ | 18 (64.3) | 10 (35.7) |  |
|  | Tanzania | <25 | 59 (42.4) | 80 (57.6) | 5.8 (0.331) |
|  |  | 25-34 | 54 (38.6) | 86 (61.4) |  |
|  |  | 35-44 | 49 (48.5) | 52 (51.5) |  |
|  |  | 45-54 | 34 (43.6) | 44 (56.4) |  |
|  |  | 55-64 | 22 (33.8) | 43 (66.2) |  |
|  |  | 65+ | 54 (36.2) | 95 (63.8) |  |
|  | Uganda | <25 | 67 (47.2) | 75 (52.8) | 5.3 (0.375) |
|  |  | 25-34 | 62 (43.1) | 82 (56.9) |  |
|  |  | 35-44 | 22 (33.8) | 43 (66.2) |  |
|  |  | 45-54 | 23 (39.7) | 35 (60.3) |  |
|  |  | 55-64 | 5 (38.5) | 8 (61.5) |  |
|  |  | 65+ | 10 (58.8) | 7 (41.2) |  |
| **Education** | Kenya | None | 7 (63.6) | 4 (36.4) | 2.9 (0.408) |
|  |  | Primary | 62 (62.6) | 37 (37.4) |  |
|  |  | Secondary | 298 (64.8) | 162 (35.2) |  |
|  |  | Higher | 186 (70.2) | 79 (29.8) |  |
|  | Tanzania | None | 42 (37.2) | 71 (62.8) | 2.4 (0.492) |
|  |  | Primary | 163 (40.8) | 237 (59.2) |  |
|  |  | Secondary | 48 (39.3) | 74 (60.7) |  |
|  |  | Higher | 19 (51.4) | 18 (48.6) |  |
|  | Uganda | None | 82 (51.9) | 76 (48.1) | 10.3 (0.016) |
|  |  | Primary | 55 (34.6) | 104 (65.4) |  |
|  |  | Secondary | 37 (45.1) | 45 (54.9) |  |
|  |  | Higher | 15 (37.5) | 25 (62.5) |  |
| **Delay in accessing care** | Kenya | Less than 2 weeks | 490 (66.3) | 249 (33.7) | 0.0 (0.986) |
|  |  | More than 2 weeks | 63 (65.6) | 33 (34.4) |  |
|  | Tanzania | Less than 2 weeks | 176 (41.1) | 252 (58.9) | 0.1 (0.712) |
|  |  | More than 2 weeks | 96 (39.3) | 148 (60.7) |  |
|  | Uganda | Less than 2 weeks | 151 (47.8) | 165 (52.2) | 9.6 (0.002) |
|  |  | More than 2 weeks | 38 (30.9) | 85 (69.1) |  |
| **Treatment-seeking steps** | Kenya | 1(straight to clinic) | 372 (64.8) | 202 (35.2) | 2.4 (0.297) |
|  |  | 2 | 153 (68.3) | 71 (31.7) |  |
|  |  | 3+ | 28 (75.7) | 9 (24.3) |  |
|  | Tanzania | 1(straight to clinic) | 91 (41.2) | 130 (58.8) | 1.7 (0.426) |
|  |  | 2 | 80 (43.7) | 103 (56.3) |  |
|  |  | 3+ | 101 (37.7) | 167 (62.3) |  |
|  | Uganda | 1(straight to clinic) | 94 (49.5) | 96 (50.5) | 5.8 (0.056) |
|  |  | 2 | 59 (39.1) | 92 (60.9) |  |
|  |  | 3+ | 36 (36.7) | 62 (63.3) |  |
| **First care accessed** | Kenya | Recruitment clinic | 372 (64.8) | 202 (35.2) | 5.2 (0.156) |
|  |  | Clinic | 98 (74.8) | 33 (25.2) |  |
|  |  | Pharmacy/drug shop | 44 (62.9) | 26 (37.1) |  |
|  |  | Self-treatment | 39 (65.0) | 21 (35.0) |  |
|  | Tanzania | Recruitment clinic | 91 (41.2) | 130 (58.8) | 4.0 (0.265) |
|  |  | Clinic | 135 (38.0) | 220 (62.0) |  |
|  |  | Pharmacy/drug shop | 29 (51.8) | 27 (48.2) |  |
|  |  | Self-treatment | 17 (42.5) | 23 (57.5) |  |
|  | Uganda | Recruitment clinic | 94 (49.5) | 96 (50.5) | 6.2 (0.101) |
|  |  | Clinic | 76 (39.4) | 117 (60.6) |  |
|  |  | Pharmacy/drug shop | 6 (31.6) | 13 (68.4) |  |
|  |  | Self-treatment | 13 (35.1) | 24 (64.9) |  |
| **Any AB use in pathway** | Kenya | No | 452 (65.5) | 238 (34.5) | 0.7 (0.388) |
|  |  | Yes | 101 (69.7) | 44 (30.3) |  |
|  | Tanzania | No | 197 (42.2) | 270 (57.8) | 1.6 (0.202) |
|  |  | Yes | 75 (36.6) | 130 (63.4) |  |
|  | Uganda | No | 132 (46.0) | 155 (54.0) | 2.6 (0.108) |
|  |  | Yes | 57 (37.5) | 95 (62.5) |  |
| TOTAL |  |  |  |  |  |
